# Projected health and economic effects of nonavalent versus bivalent human papillomavirus vaccination in preadolescence in the Netherlands

**DOI:** 10.1101/2023.12.27.23300574

**Authors:** Birgit Sollie, Johannes Berkhof, Johannes A. Bogaards

## Abstract

**Background:** Most European countries offer human papillomavirus (HPV) vaccination through organized immunisation programmes, but the choice of vaccine varies. We compared the expected health and economic effects of the currently used bivalent vaccine, targeting HPV-16/18, and the nonavalent vaccine, targeting seven additional genotypes, for the Netherlands.

**Methods:** We estimated the incremental impact of nonavalent versus bivalent vaccination in a cohort of 100,000 girls and 100,000 boys offered vaccination at age 10, by projecting type-specific infection risk reductions onto expected number of cervical screening outcomes, HPV-related cancers, and treatments for anogenital warts and recurrent respiratory papillomatosis (RRP). In the base-case, we assumed two-dose vaccination with 60% uptake, lifelong partial cross-protection against HPV-31/33/45 for the bivalent vaccine and EUR 25 extra costs per dose for the nonavalent vaccine. Cost-effectiveness was assessed by comparing the incremental cost-effectiveness ratio (ICER) per life-year gained (LYG) with the Dutch threshold of EUR 20,000/LYG.

**Findings:** Compared with bivalent vaccination, nonavalent vaccination prevents an additional 1320 high-grade cervical lesions, 70 cancers, 34,000 anogenital warts episodes and 30 RRPs; and generates EUR 4·0 million discounted savings from fewer treatments. The ICER is EUR 6192 (95% credible interval: 4166; 7916)/LYG in the base-case, but exceeds the cost-effectiveness threshold when cross-protection for the bivalent vaccine extends to non-31/33/45 genotypes or when vaccine efficacy wanes past age 20 with either vaccine.

**Interpretation:** Sex-neutral vaccination with the nonavalent vaccine is likely to be cost-effective. Long-term monitoring of type-specific vaccine effectiveness is essential because of the impact of cross-protection and waning efficacy on cost-effectiveness.

## Introduction

The sexually transmitted human papillomavirus (HPV) can cause a variety of diseases in the anogenital and oropharyngeal body sites, predominantly cervical cancer [1]. Most European countries offer free-of-charge vaccination against HPV through organized immunisation programmes. However, the type of vaccine offered varies across Europe.

There are currently three prophylactic HPV vaccines authorized for use in both males and females in the European Union (EU) (see https://www.ema.europa.eu/en/homepage for an European public assessment report (EPAR) on each product): a bivalent vaccine (Cervarix®, GlaxoSmithKline Biologicals SA) targeting HPV genotypes 16 and 18, which are associated with approximately 70% of cervical cancers and the majority of other HPV-related cancers; a quadrivalent vaccine (Gardasil®, Merck Sharp & Dohme BV) targeting HPV-16 and −18 as well as low-risk (LR) HPV genotypes 6 and 11, which are associated with anogenital warts and recurrent respiratory papillomatosis (RRP); and, since 2015, a nonavalent vaccine (Gardasil9®, Merck Sharp & Dohme BV), targeting an additional five high-risk (HR) HPV genotypes (31, 33, 45, 52 and 58) which are found in high-grade precancerous cervical lesions and cancer.

The prices of the bivalent and quadrivalent HPV vaccines have strongly declined since market introduction because of tender-based procurement [2]. Recent additions to the EU Tenders Electronic Daily website (https://ted.europa.eu/) show that the nonavalent HPV (9vHPV) vaccine is now the most often procured for use in national or regional immunisation programmes. Yet a number of countries―including Bulgaria, Czechia, Finland, Norway and the Netherlands―still use the bivalent (2vHPV) or quadrivalent (4vHPV) vaccine, mainly because its price is considerably lower than that of the nonavalent vaccine.

Most countries that implemented nonavalent vaccination used the 4vHPV vaccine before, in which case cost-effectiveness is determined solely by weighing the extra cost of the 9vHPV vaccine against the extra protection against the five additional HR HPV types. However, unlike the 4vHPV vaccine, it is widely recognized that the 2vHPV vaccine provides cross-protection against genotypes phylogenetically related to HPV-16 or −18, in particular HPV-31, −33, and −45 [3–7], which should be taken into account when evaluating the additional benefits from 9vHPV vaccination. Dynamic modelling studies that compared the expected health and economic effects from 9vHPV versus 2vHPV vaccination in high-income countries arrived at different conclusions regarding the cost-effectiveness of the 9vHPV vaccine [8–16]. The comparison is particularly challenging because it requires thoughtful consideration about the benefits of preventing diseases associated with LR HPV types, as these may have different weights in decision-making. Moreover, both vaccines are produced by different companies and vaccination costs are subject to competitive bidding [2], which should be reflected in realistic price differences in the health economic evaluation.

The purpose of this study is to provide a comprehensive comparison of the 9vHPV and 2vHPV vaccines in the Dutch setting of sex-neutral vaccination with tender-based procurement. We used a data-driven approach in which the direct and indirect effects of vaccination were projected onto the occurrence of all HPV-associated diseases, including the outcomes of primary HPV-based screening for cervical cancer. In sensitivity analyses, we consider several scenarios related to vaccine efficacy (including cross-protection and waning), vaccine uptake and expected price differences between the two vaccines. In cost-effectiveness analyses, we include all cost savings, but focus primarily on health gains from cancer prevention.

## Methods

Our assessment builds upon the evidence synthesis framework that we previously developed to estimate the health and economic impact of sex-neutral compared to girls-only HPV vaccination [17–19]. This framework allows for lifetime evaluation of an HPV-naive birth cohort in terms of HPV-associated disease occurrence and medical costs incurred and, by applying Bayesian analyses to life tables, yields credible intervals for the relevant outcomes. To compare the 9vHPV and 2vHPV vaccines in the setting of sex-neutral vaccination, we simulated a cohort of girls and boys invited for HPV vaccination at the target vaccination age of 10 years in the national immunisation programme in the Netherlands. We estimated the total health and economic effects under vaccination with either the 9vHPV or 2vHPV vaccine for this hypothetical cohort with respect to the following events: colposcopy referral and detected precancerous lesion within the cervical screening programme, diagnosis of cervical cancer as well as HPV-induced oropharyngeal, anal, vulvar, vaginal or penile cancer, treatment for anogenital warts and onset of recurrent respiratory papillomatosis (RRP).

Our data-driven approach can be divided into three steps. First, we estimated the expected number of events in the hypothetical cohort in the absence of HPV vaccination. Second, we estimated how many events will be prevented under 9vHPV or 2vHPV vaccination, by projecting type-specific HPV infection risk reductions onto numbers of type-specific vaccine-preventable diseases. For this projection, we developed a statistical model that describes the onset age distribution of the causal HPV infection in subjects with HPV-associated cancer. Third, we translated the difference in health and economic effects between the two vaccines into an incremental cost-effectiveness ratio (ICER) of 9vHPV versus 2vHPV vaccination, conditional on assumptions about long-term vaccine efficacy against vaccine-targeted and cross-protected genotypes, vaccine uptake, and costs. A detailed description is given in the Supplementary Annex A, and is summarized below.

### Expected number of events in the absence of HPV vaccination

To estimate the number of expected events in the absence of HPV vaccination, we used population-level data on the age-specific incidence of HPV-associated cancers, RRPs, and anogenital warts, and the detection rate of cervical cancer screening outcomes in the Netherlands. We assumed that HPV vaccination effects on vaccine-preventable diseases were not yet measurable in the Netherlands until 2020. This is plausible because 2vHPV vaccination is assumed to have no effect on LR HPV genotypes, and HPV-vaccinated women were not eligible for screening in the Netherlands until 2023.

To estimate the detection rate of high-grade precancerous lesions through HPV-based screening, we analysed the outcomes of the Dutch cervical screening programme between 2017-2019. The expected number of colposcopies per screening round was computed from the expected number of cervical intraepithelial neoplasia (CIN) grade 2/3 diagnoses by multiplying the latter with the number of colposcopies needed to detect one precancerous lesion, stratified by screening round [20]. The age-specific incidence and survival rates for cervical cancer and the other HPV-related cancers were estimated from Netherlands Cancer Registry data over the years 2015-2019. The rate of anogenital warts episodes was obtained from the national surveillance of sexually transmitted infections, combined with a GP registry study. We used international publications to obtain the age-specific incidence of RRP. We made a distinction between adult-onset RRP, resulting from a self-acquired HPV infection, and juvenile-onset RRP, due to mother-to-child HPV transmission during childbirth. Only the expected future children of the girls in the hypothetical cohort were considered at risk for juvenile-onset RRP. For RRP patients, we assumed an exponentially distributed duration of the disease with a mean of 10 years. Sources and details on calculation for warts and RRP incidence are given in Supplementary Annex A. Life expectancy of the cohort was based on recent life-tables collected from Statistics Netherlands (https://www.cbs.nl/en).

In estimating age-specific event rates, we took into account the uncertainty of the data by applying a Bayesian analysis. We ran 1000 simulations in which the parameters were sampled from posterior distributions, informed by data and non-informative priors (see Supplementary Annex A for details). The outcomes are reported in terms of 95% credible intervals (CI), containing the 2.5th and 97.5th percentiles of the results obtained via simulation.

### Expected number of events prevented by HPV vaccination

The expected number of events prevented by HPV vaccination in the simulated cohort was computed for each specific vaccination scenario. To this end, we first estimated the event-specific attribution to HPV genotypes of interest, i.e. those to which the 9vHPV or 2vHPV vaccine provide protection. The HPV genotype attribution of precancerous lesions was estimated from Dutch screening trial data using a previously developed maximum likelihood method [21]. HPV genotype attributions for HPV-associated cancers, warts and RRP were obtained from the literature (see Supplementary Annex A for references). Next, we projected age- and type-specific risk reductions from vaccination onto the expected number of events in the absence of vaccination, under specific scenarios regarding long-term vaccine efficacy and vaccine uptake (see paragraph “Vaccine uptake and efficacy”). HPV infection risk reductions for all relevant HR HPV genotypes were obtained from a previously developed model for heterosexual type-specific HPV transmission [22]. We assumed that the simulated cohort experiences age-specific infection risks that apply to the post-vaccination equilibrium, an assumption that we showed to be valid after approximately 10 years of HPV vaccination [19].

To calculate the number of diagnosed high-grade cervical lesions (CIN2/3) averted, reductions in type-specific HPV prevalence at each screening round were projected onto the number of expected CIN2/3 diagnoses attributed to these types. The number of colposcopies averted was computed by recalculating the number of colposcopies needed to detect one precancerous lesion, taking into account the reduced CIN2/3 risks in HPV-positive women with abnormal cytology [20]. To translate type-specific HPV infection incidence reductions into cancer risk reductions, we estimated the period from HPV infection to cancer diagnosis for each of the six cancers included in our analysis (see Supplementary Annex A). The risk reductions for LR HPV genotypes could not be obtained from our HPV transmission model, as the model was only calibrated to HR HPV genotypes. However, there is strong evidence that the herd effects for the LR HPV genotypes are large [23], presumably at least as large as the herd effects for HPV-18 [24]. We therefore used the average reduction in HPV-18 prevalence to approximate the herd effects for HPV-6 and −11. Uncertainty in the differential impact of HPV vaccines primarily follows from uncertainty in HPV genotype attributions to the events of interest, and these were incorporated via a Bayesian analysis.

### Incremental cost-effectiveness analysis

We conducted a health economic analysis from a societal perspective, in which we considered all medical and non-medical costs related to HPV-related diseases. Cost of medical procedures related to the events of interest (indexed to the year 2023 via the consumer price index) are listed in Table 1. For 2vHPV vaccination at age 10 years, we assumed a total vaccination cost of EUR 65 per person using a two-dose vaccination schedule, as previously reported for the Netherlands [18]. The total vaccination cost for 9vHPV vaccination was set at EUR 115 per person in the base-case scenario, based on an anticipated price difference of EUR 25 per dose, i.e. EUR 50 for a two-dose schedule, between the 9vHPV and 2vHPV vaccines [2].

**Table 1.**
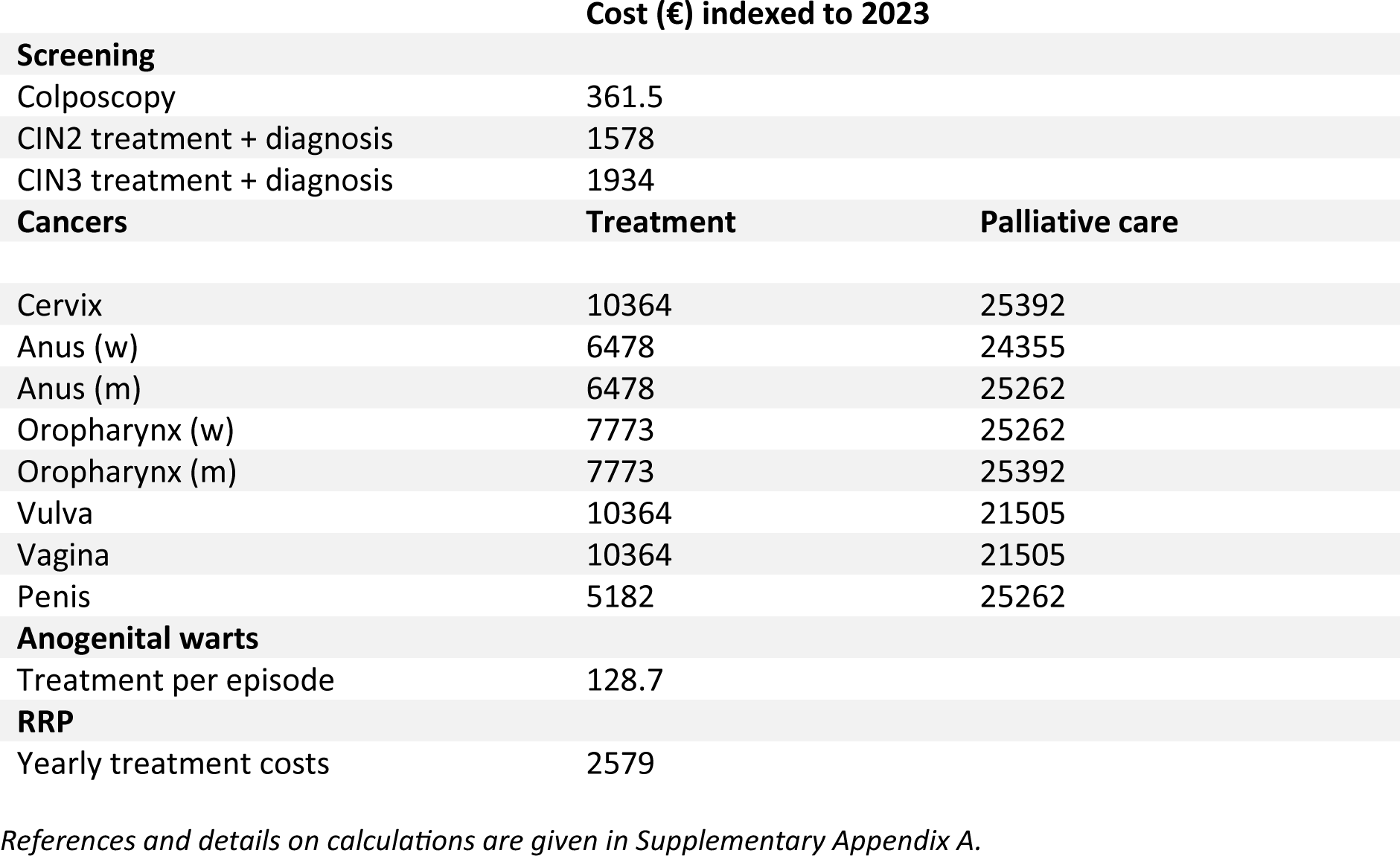
Assumed costs (in €) indexed to the year 2023 using the Consumer Price Index.

Events expected to be prevented by HPV vaccination were translated into cost savings and life-years gained for each specific vaccination scenario. The number of life-years gained by preventing cancer cases was calculated using cancer survival data collected from the Netherlands Cancer Registry (https://iknl.nl/en), in combination with data on overall survival from Statistics Netherlands. Future costs and effects were discounted by 3% and 1.5% per year, respectively, according to current Dutch guidelines. We then computed an incremental cost-effectiveness ratio (ICER) for each particular comparison of vaccination with the 9vHPV versus 2vHPV vaccine, which is the ratio of the difference in discounted costs and the difference in discounted life-years gained (LYG). Switching to 9vHPV vaccination was considered cost-effective when the ICER was below the Dutch threshold for preventive interventions of EUR 20,000 per (quality-adjusted) LYG.

This study adheres to HPV-FRAME, a quality framework for the reporting of mathematical modelling evaluations of HPV-related cancer control. The checklist is reported in Supplementary Annex C.

### Vaccine uptake and efficacy

In the base-case scenario, we assumed 60% vaccine uptake in line with average figures in recently vaccinated preadolescent cohorts (https://www.vzinfo.nl/prestatie-indicatoren/vaccinatiegraad-hpv). In sensitivity analyses, we also considered 50% (historic average) and 70% (optimistic uptake). Vaccine uptake among boys was set equal to that among girls in all scenarios.

Vaccine efficacy (VE) against HPV genotypes 16 and 18 in our analysis was set to a pooled estimate of 98% in per-protocol populations of 2vHPV and 4vHPV vaccine trials with endpoints of HPV-16/18-associated CIN2/3 [17]. Although VE estimates for non-cervical sites are less precise, we conjectured the same genotype-specific efficacy for all sites. In addition, we assumed that the 9vHPV vaccine has the same VE against diseases caused by non-16/18 vaccine genotypes as against those caused by HPV-16/18 [25]. In our base-case scenario we further assumed lifelong partial cross-protection for the 2vHPV vaccine against genotypes 31, 33 and 45. The protective effect against these HR HPV genotypes has been consistently demonstrated in trials with the 2vHPV vaccine [3], as well as in post-vaccination surveillance Scotland and Finland [4, 6] as well as in the Netherlands [5, 7] with up to 10 years follow-up. We assumed cross-protective vaccine efficacies of 75%, 50% and 80% against HPV-31, −33 and −45, respectively, in the base-case scenario. We also considered alternative scenarios in sensitivity analysis, with VE assumptions for the HR HPV genotypes given in Table 2.

**Table 2.** Vaccine efficacies for the high-risk HPV genotypes.

|  | 16 | 18 | 31 | 33 | 35 | 39 | 45 | 51 | 52 | 58 |
| --- | --- | --- | --- | --- | --- | --- | --- | --- | --- | --- |
| **Base-case** |  |  |  |  |  |  |  |  |  |  |
| 2vHPV | 98% | 98% | 75% | 50% |  |  | 80% |  |  |  |
| 9vHPV | 98% | 98% | 97% | 97% |  |  | 97% |  | 97% | 97% |
| **Sensitivity analysis** |  |  |  |  |  |  |  |  |  |  |
| 2vHPV no cross-protection | 98% | 98% |  |  |  |  |  |  |  |  |
| 2vHPV high cross-protection | 98% | 98% | 66% | 41% | 40% |  | 81% |  | 36% | 30% |
| 2vHPV very high cross-protection | 98% | 98% | 88% | 68% |  | 75% | 82% | 54% |  |  |

*The base-case scenario considers values that are between the 95% confidence bounds of various empirical estimates across a range of settings [3–7]. In the most conservative scenario, we only assume protection from 2vHPV against vaccine types 16 and 18. In a more liberal scenario, we considered cross-protection against phylogenetically related genotypes, for which cross-protection has been demonstrated in the Netherlands [5]. In the most extreme scenario, we considered cross-protective efficacies at the level reported in the EMA EPAR documentation of the 2vHPV vaccine.*

### Sensitivity analysis

In addition to vaccine uptake and degree of cross-protection from the 2vHPV vaccine, we analysed the influence of the degree of protection against LR HPV genotypes, the degree of waning efficacy and vaccine price difference. We considered two extreme scenarios regarding the protection against LR HPV genotypes; one in which the LR HPV genotypes are excluded from the analysis, and one in which we assumed complete elimination of anogenital warts and RRP due to herd immunity from vaccination with the 9vHPV vaccine. Regarding waning efficacy, we considered a waning scenario for all non-16/18 genotypes included in the analysis where waning starts at age 20 and the efficacy decreases at a constant rate, leaving only 5% of initial efficacy at age 40. We assume that efficacy against non-16/18 HR HPV infections would be maintained for at least 10 years after vaccination, in line with recent data on the long-term effectiveness of both the 9vHPV and 2vHPV vaccines [26, 27]. The difference in costs of vaccination were varied between EUR 35 and EUR 70 for a two-dose schedule, in accordance with the uncertainty range in a multivariable analysis of procurement data across European countries [2].

Furthermore, we expressed the ICER in terms of incremental cost per quality-adjusted life-years gained (QALYG), taking into account the loss in quality of life from non-lethal conditions, i.e. CIN2/3 diagnoses, anogenital warts and RRP (see Supplementary Annex for specific utilities used). Finally, to accommodate international comparisons, we considered a scenario where costs and effects were both discounted by 3% per year as recommended by the WHO, and with the threshold for cost-effective intervention set at EUR 50,000 per LYG, close to the Dutch GDP per capita.

We applied both one-way and two-way sensitivity analyses. We only present two-way sensitivity analyses for combinations of one-way analyses with ICERs higher than in the base case but below the cost-effectiveness threshold (to see if their combination could lead to an ICER above the threshold); or with one ICER above the threshold and the other below the base-case ICER (to see if their combination still produces an ICER above the threshold).

## Results

### Base-case scenario

In a cohort of 100,000 girls and 100,000 boys, a total of 17,765 colposcopies, 8290 CIN2/3 diagnoses, 710 cervical cancer cases and 585 other HPV-associated cancers are expected without HPV vaccination. Vaccination with the 2vHPV vaccine is expected to avert 10,895 colposcopies, 5440 related CIN2/3 diagnoses, 575 cervical cancer cases and 500 of the other cancers through a combination of direct protection and herd immunity with 60% vaccine uptake. Vaccination with the 9vHPV vaccine is expected to avert an additional 2440 colposcopies, 1320 CIN2/3 diagnoses, 45 cervical cancer cases and 25 other cancers (Fig. 1).

**Figure 1:**
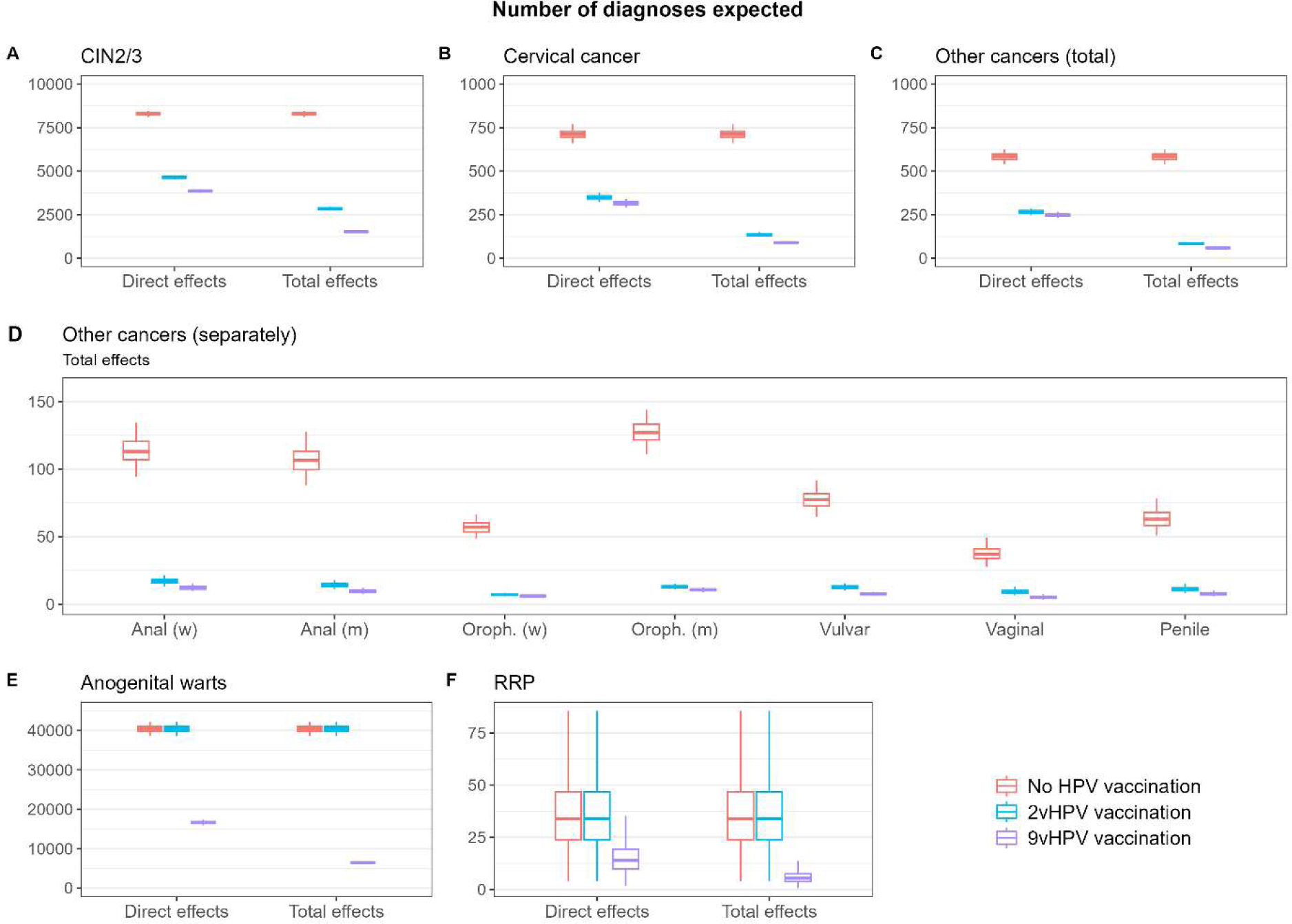
Expected number of events per cohort of 100,000 girls and 100,000 boys. Number of CIN2/3 diagnoses expected (panel A), cervical cancer cases expected (panel B), other cancer cases expected (panel C), other cancer cases expected per site (panel D), anogenital warts episodes expected (panel E) and RRP patients expected (panel F) in case of no vaccination (red), vaccination with the 2v vaccine (blue) and vaccination with the 9v vaccine (purple). Panels A-C and E-F show the direct effects only and total effects, consisting of direct plus herd protection. Panel D only shows the results for direct effects plus herd effects. Boxplots display median and interquartile range of predictions, with whiskers denoting the 95% credible intervals.

For the non-cancer outcomes associated with HPV-6 and −11, approximately 40,000 anogenital warts episodes and 34 RRP patients per cohort of 100,000 girls and 100,000 boys, including 8 cases of juvenile-onset RRP, are expected without HPV vaccination. These diseases cannot be prevented by 2vHPV vaccination, but we estimate that, through a combination of direct protection and herd immunity, 9vHPV vaccination at 60% vaccine uptake will prevent approximately 34,000 anogenital warts episodes and 30 RRPs (Fig. 1).

The expected number of prevented events can be translated into LYG and monetary savings. At 60% vaccine uptake, sex-neutral vaccination with the 2vHPV vaccine leads to a gain of 6·7 thousand life-years (3·5 thousand discounted) per 100,000 girls and 100,000 boys, and saves a total of EUR 30·4 million (EUR 9·8 million discounted) by averting colposcopies, CIN2/3 diagnoses and HPV-related cancers. Vaccination with the 9vHPV vaccine provides an additional gain of 435 life-years (224 discounted) and additional savings of EUR 9·2 million (EUR 4·0 million discounted), the latter mainly through prevention of warts and RRP and reduced diagnosis of CIN2/3. Figure 2 shows the LYG and costs through HPV vaccination, both for 9vHPV and 2vHPV vaccination, broken down by type of HPV-associated disease. Most of the monetary savings come from reduced diagnosis of CIN2/3. Savings from the prevention of anogenital warts and RRP are lowest in absolute terms, but after 3% discounting per year for future effects, they become comparable to monetary savings from the prevention of cervical cancer.

**Figure 2:**
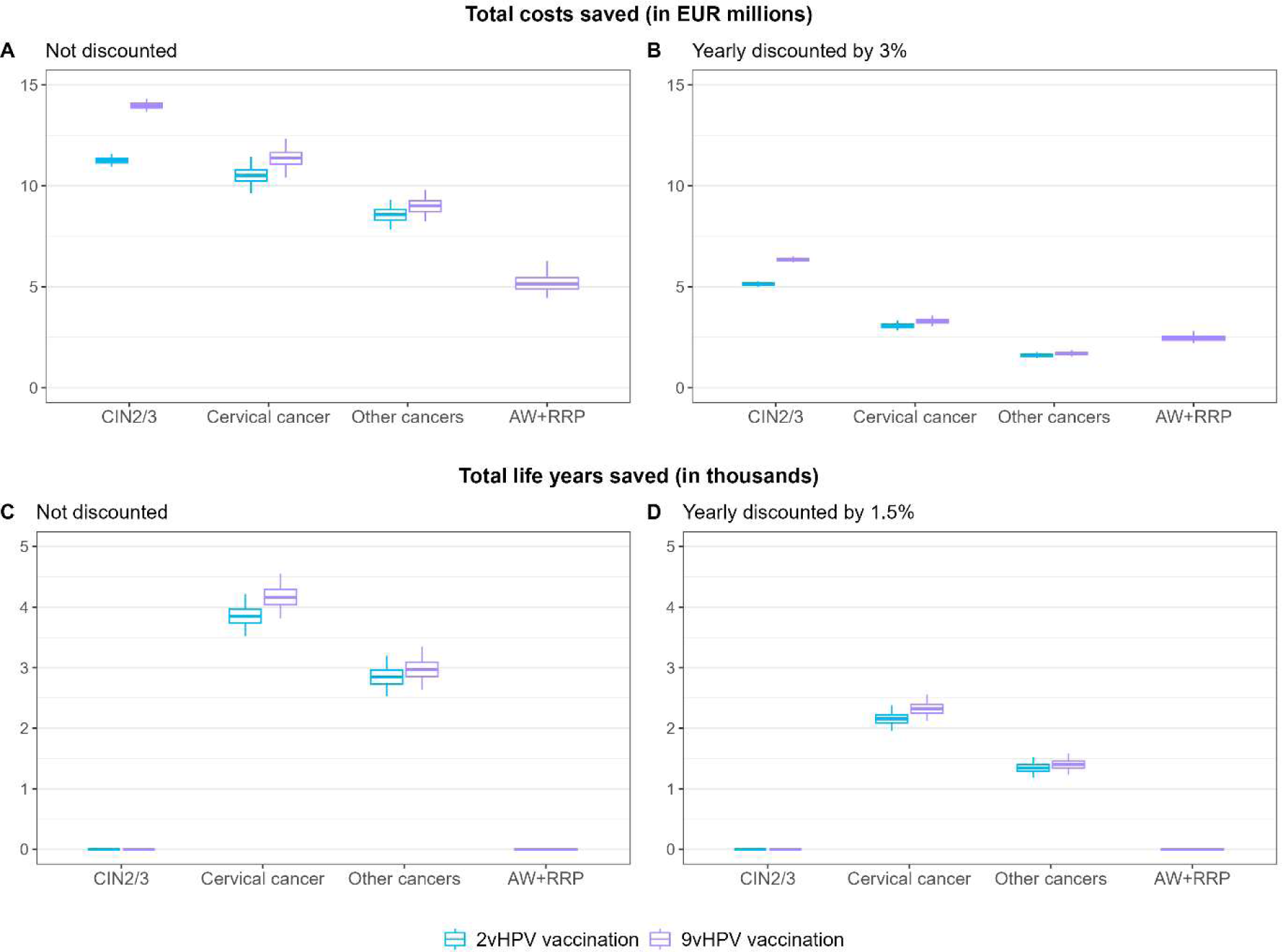
Expected health and economic effects per cohort of 100,000 girls and 100,000 boys. Health and economic effects of HPV vaccination with 2vHPV vaccine (blue) or 9vHPV vaccine (purple), either not discounted (left) or discounted (right). Boxplots display median and interquartile range of predictions, with whiskers denoting the 95% credible intervals.

The ICER of 9vHPV versus 2vHPV vaccination at annual discount rates of 1.5% for effects and 3% for costs is EUR 6192 (95% CI: 4166; 7916) per LYG and lies below the cost-effectiveness threshold for preventive interventions of EUR 20,000/LYG.

### Sensitivity analysis

Our conclusion remain similar under discounting of 3% per year for both effects and costs, with corresponding threshold for cost-effective interventions of EUR 50,000 per LYG (Supplementary Annex B).

Vaccination with the 9vHPV instead of 2vHPV vaccine remains cost-effective in almost all scenarios investigated in one-way sensitivity analyses (Fig. 3). Switching from 2vHPV to 9vHPV vaccination remains cost-effective if the effect of the 9vHPV vaccine on LR HPV genotypes is ignored and would even lower the costs of the programme if cross-protection to non-16/18 genotypes from the 2vHPV vaccine is ignored. However, the cost-effectiveness profile may change when increasing the degree of cross-protection assumed for the 2vHPV vaccine. Increasing the degree of cross-protection for the 2vHPV vaccine by extending cross-protection to HPV-35/52/58, i.e. genotypes with close phylogenetic relationship to HPV-16 or −18 [5], leads to an ICER close to the threshold of EUR 20,000 per LYG. When assuming very high cross-protection to HPV-31/33/45 as well as to HPV-39/51 at levels reported in EMA-EPAR documentation, the switch to 9vHPV vaccination is no longer cost-effective. The ICER under increased cross-protection from the 2vHPV vaccine remains above the threshold when combined with several other scenarios that yield a lower ICER compared to the base-case in one-way analyses (see Supplementary Annex B).

**Figure 3:**
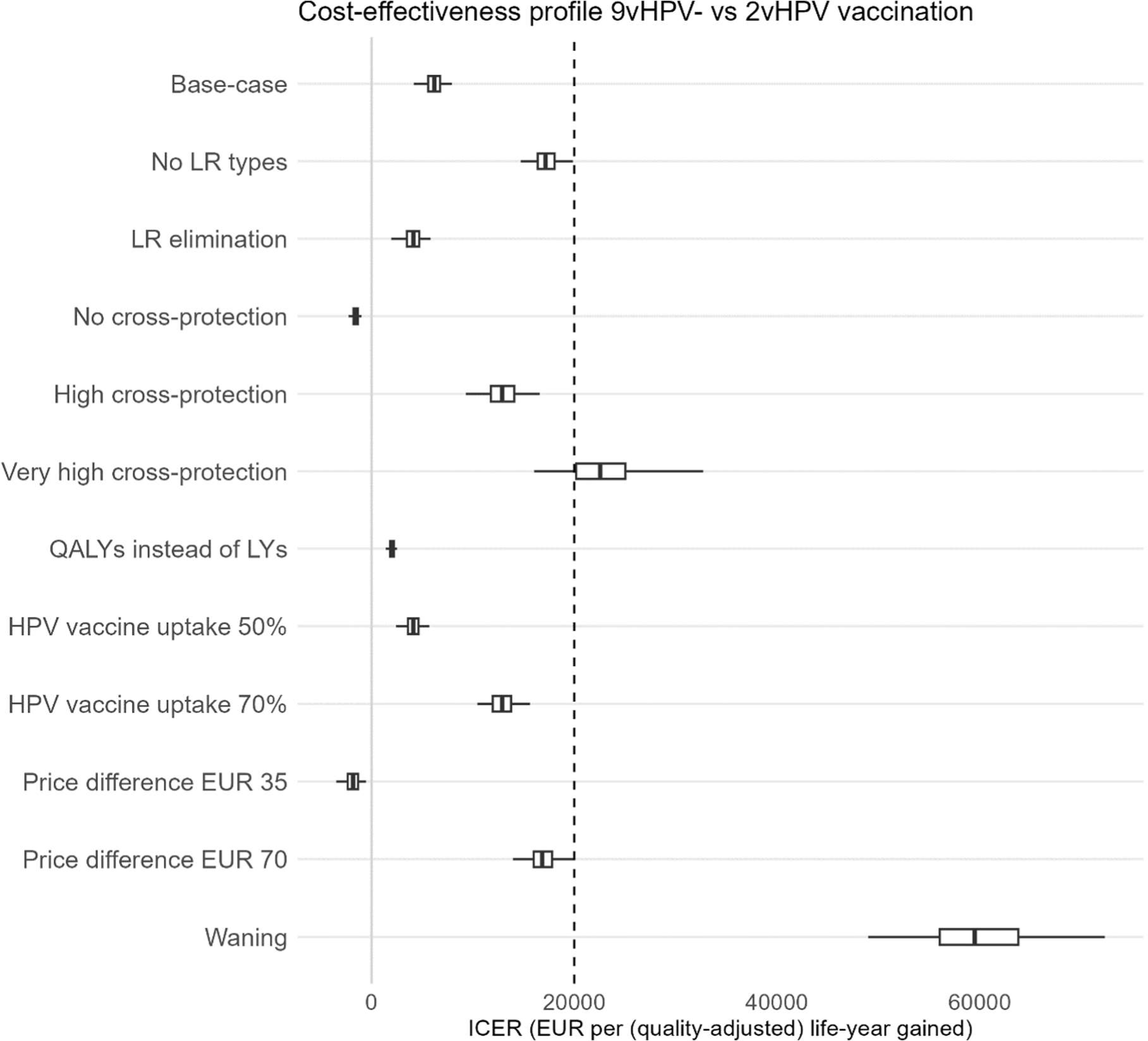
Incremental cost-effectiveness ratio (ICER) by scenario in one-way sensitivity analyses. ICERs of 9vHPV versus 2vHPV vaccination for different scenarios with respect to low-risk (LR) HPV genotypes, cross-protection from 2vHPV vaccine, inclusion of quality-adjusted life-years (QALYs), vaccine uptake, expected price differences between the 9vHPV and 2vHPV vaccines for a two-dose vaccination schedule, and waning efficacy for non-16/18 high-risk HPV genotypes. The light-grey vertical line corresponds to an ICER equal to zero. The cost-effectiveness threshold of €20,000 per (quality-adjusted) life-year gained is displayed by the dashed vertical line. All scenarios with an ICER to the left of this line support the conclusion that the 9vHPV vaccine is cost-effective compared to the 2vHPV vaccine. Boxplots display median and interquartile range of predictions, with whiskers denoting the 95% credible intervals.

Replacing LYG by QALYG in the analysis increases the benefit of 9vHPV vaccination and leads to a lower ICER compared to the base-case scenario. Using QALYG instead of LYG also leads to an ICER below the cost-effectiveness threshold in all scenarios with increased cross-protection for the 2vHPV vaccine (Supplementary Annex B). A decreased vaccine uptake leads to a decrease in ICER, whereas an increased vaccine uptake leads to an increase in ICER, but the ICER remains below the cost-effectiveness threshold at 70% vaccine uptake, unless LR HPV genotypes are excluded from the analysis (Supplementary Annex B). If the price difference between the two vaccines is decreased from EUR 50 to EUR 35 for a two-dose schedule, switching from 2vHPV to 9vHPV vaccination will lower the costs of the programme. If the price difference is increased to EUR 70 for a two-dose schedule, the ICER remains below the threshold of EUR 20,000 per LYG, unless LR HPV genotypes are excluded or vaccine uptake is 70% (Supplementary Annex B).

The assumption of waning efficacy from age 20 for all non-16/18 genotypes had the greatest impact on the ICER. At a constant waning rate, where protection was virtually lost by age 40, it would cost almost EUR 60,000 (95% CI: 48,995; 72,315) per LYG to switch from 2vHPV to 9vHPV vaccination. The assumption of waning efficacy also pushes ICERs above the cost-effectiveness threshold in two-way sensitivity analyses, unless the ICER is based on QALYG instead of LYG (Supplementary Annex B).

## Discussion

In this study, we compared the projected health and economic effects of HPV vaccination in the Dutch national immunisation programme under sex-neutral vaccination with either the 9vHPV or 2vHPV vaccine. Our results suggest that switching from 2vHPV to 9vHPV vaccination is likely to be cost-effective according to criteria for preventive interventions in the Netherlands.

In our base-case scenario, assuming 60% vaccine uptake at age 10 and lifelong protection against vaccine-targeted and cross-protected HPV types, 9vHPV vaccination has a favourable cost-effectiveness ratio compared to 2vHPV vaccination of EUR 6192 per LYG at an additional two-dose vaccination cost for the 9vHPV vaccine of EUR 50. This favourable cost-effectiveness ratio was retained under several scenarios for vaccine uptake and price difference. However, the cost-effectiveness of 9vHPV vaccination is less clear when the 2vHPV vaccine provides lifelong cross-protection against a wider range of HR HPV genotypes than is typically assumed, or when efficacy against non-16/18 HR HPV genotypes would start to decline 10 years after vaccination with either vaccine. Conversely, if cross-protection is ignored, or the 9vHPV vaccine costs only EUR 35 more per two-dose schedule, switching to 9vHPV vaccination would even reduce the costs of the programme.

Our analysis pinpoints the importance of durable cross-protection when assessing the incremental benefit of vaccination with the 9vHPV versus 2vHPV vaccine. Estimates of cross-protective efficacy are still uncertain and seem to vary by setting. Our base-case scenario assumed cross-protective efficacies against HPV-31, −33 and −45 that are consistent with empirical data across a range of settings [3–7]. We also considered a scenario of broader cross-protection, as suggested by an analysis of vaccine efficacy in relation to phylogenetic distance from HPV-16 or −18, derived from the Netherlands [5]. We could have considered more variation in cross-protection scenarios, but instead we chose to explore two extreme scenarios: one with no cross-protection to non-16/18 HPV types, and one with high and broad cross-protection, as reported in EMA-EPAR documentation. The latter scenario included cross-protection against HPV-39 and −51, even though cross-protection against (persistent) infection with these types has not been confirmed in post-vaccine surveillance studies [5–7]. We think that a scenario of no cross-protection is not relevant for the Netherlands given the context-specific data, but results under this scenario highlight the importance of cross-protection and also make our results comparable to other studies.

Dynamic modelling studies that did not account for cross-protection invariably concluded that 9vHPV vaccination would be cost-effective or even less expensive than 2vHPV vaccination, both in girls-only and in sex-neutral vaccination programmes [9–12]. In dynamic modelling studies that did allow for cross-protection, conclusions were less straightforward [8, 13–16]. However, most of these only included cervical disease outcomes [8, 13, 14], and did not consider additional gains from preventing non-cervical cancers, warts and papillomatosis. Including non-cancer outcomes would have resulted in a more favourable assessment of 9vHPV versus 2vHPV vaccination, yet health authorities may choose to disregard these gains given the imbalance in disease severity between cancers and warts and the high priority of cervical cancer prevention in many countries. However, we believe that direct medical savings resulting from an intervention should be included in economic evaluations from a healthcare payer or societal perspective. For this reason, we did incorporate all cost savings from the additional prevention of warts and papillomatosis by 9vHPV vaccination in our base-case scenario. Leaving these out still gave a favourable cost-effectiveness profile at 60% vaccine uptake, but no longer at 70% vaccine uptake.

Our analysis considered the benefit of broadening protection against HR HPV types in the context of cervical screening via primary HR HPV testing. The switch to HPV-based screening in the Netherlands in 2017 has resulted in a substantial increase in number of colposcopy referrals and CIN2/3 diagnoses [28]. As the proportion of non-16/18 HPV types in screen-positives and CIN2/3 is higher than in cervical cancer [21], the increased protection offered by the 9vHPV vaccine should yield substantial savings in HPV-based screening. Indeed, we found that the incremental cost savings from broader protection against HR HPV types were driven by the decrease in colposcopies and CIN2/3 treatments rather than by the extra numbers of cancers averted. In fact, the discounted cost savings from 9vHPV compared to 2vHPV vaccination in HPV-based screening would still exceed those from preventing warts and RRP. However, it remains to be seen whether HPV-based screening at five-year intervals is itself cost-effective in cohorts eligible for 9vHPV vaccination.

To date, only two other dynamic modelling studies have directly compared 9vHPV and 2vHPV vaccination in the context of sex-neutral vaccination programmes, taking into account all HPV-related diseases and HPV-based cervical screening outcomes [15, 16]. In these studies, situated in Norway and in the Netherlands, the investigators assumed that 2vHPV vaccination provided cross-protection against HPV-31, −33, and −45, either in the base-case or in sensitivity analysis. However, while the protection provided by 9vHPV vaccination was assumed to be lifelong, cross-protection was assumed to wane at a rate between 10-30% per year. In such a fast-waning scenario, only 3-33% of the initial cross-protective efficacy administered at age 10 would persist at age 20, which is clearly at odds with observations of sustained cross-protective efficacy for at least 10 years after 2vHPV vaccination in the Netherlands [26]. For this reason, we only considered scenarios of waning efficacy starting from the age of 20, i.e. 10 ten years after vaccination. Of note, 9vHPV vaccination has also demonstrated sustained immunogenicity and efficacy up to 10 years after vaccination of preadolescents [27]. We did not consider a scenario of waning efficacy for the 2vHPV vaccine only and lifelong efficacy for the nonavalent vaccine, which would clearly favour the 9vHPV vaccine.

Our data-driven approach differs somewhat from other dynamic modelling studies that have aimed to project the long-term impact of HPV vaccination, including our own modelling studies in the Dutch setting. The main difference with our previous hybrid modelling framework is that we did not use a microsimulation model for carcinogenesis because the differential impact of vaccination with either the 9vHPV or 2vHPV vaccine on HR HPV genotypes is driven by types, notably HPV-52 and −58, for which a detailed model specification in terms of natural disease progression is still challenging. We are much more certain about their specific attribution to CIN2/3 and cancer, based on well-established statistical methods for estimating genotype attribution in precancerous lesions [21] and for estimating the time from HPV infection to cancer diagnosis [29]. Consequently, we developed a work-around based on projections of type-specific infection risk reductions onto the expected CIN2/3 and cancer diagnoses attributed to these genotypes.

This study has some notable limitations. First, our analysis relied on recent population-level data but ignored the upward trend in the HPV-related disease burden over the past decades. If this trend would continue in the absence of HPV vaccination, our estimates of the impact of HPV vaccination are likely to be on the conservative side, and the same holds for the incremental benefit of 9vHPV as compared to 2vHPV vaccination. Second, we did not account for the clustering of anal HPV infections and anal cancer in men who have sex with men (MSM). On the basis of a recently developed model for homosexual transmission of HPV 16, we concluded that the relative reduction in anal HPV 16 prevalence among MSM from vaccinating boys in preadolescence are comparable to relative reductions in genital HPV 16 infections among heterosexuals [30]. Hence, projecting relative reductions from heterosexual transmission onto expected number of anal cancers might be considered reasonable for MSM too. Third, in the absence of a calibrated transmission model for HPV-6 and −11, we made a conservative assumption about the herd effects for the LR HPV genotypes covered by the 9vHPV vaccine, and also considered an extreme scenario in which the LR HPV genotypes were eliminated. Finally, the cost savings in HPV-based screening may have been slightly underestimated, because we did not account for a possible reduction in the demand for cytological triage and repeat tests after a positive HPV result. Nevertheless, the cost savings in HPV-based screening will be mainly determined by the reduction in referrals for colposcopy and CIN2/3 diagnoses, which have been captured accordingly.

In conclusion, sex-neutral vaccination with the 9vHPV vaccine is likely to be cost-effective compared to 2vHPV vaccination within the national immunization programme of the Netherlands. Whether the ICER remains below the Dutch cost-effectiveness threshold for preventive interventions is mainly determined by the presumed breadth of protection provided by the 2vHPV vaccine. It is therefore advisable to monitor the long-term effectiveness of the 2vHPV vaccine on a type-specific level, and to update projections on the impact and cost-effectiveness of HPV vaccination when new data become available. The influx of HPV-vaccinated birth cohorts into the Dutch cervical cancer screening programme will provide unique insights in this respect, that can be used to reevaluate the relative merit of 9vHPV vaccination.

## Data Availability

Data and code are available through GitHub, via BirgitSollie/HPV-COMPARE

https://github.com/BirgitSollie/HPV-COMPARE

## Acknowledgements

We thank emeritus professor Frederik G. Dikkers for the use of RRP patient cost data, and emeritus professors Chris J.L.M. Meijer and Gemma G. Kenter, as well as various stakeholders, for helpful discussions on the comprehensive comparison of nonavalent and bivalent HPV vaccines within the context of HPV-related disease prevention in the Netherlands.

