## Supplementary Material for "Projected health and economic effects of nonavalent versus bivalent human papillomavirus vaccination in preadolescence in the Netherlands"

Birgit Sollie, Johannes Berkhof, Johannes A. Bogaards
Amsterdam UMC, Dept. Epidemiology & Data Science, Amsterdam, The Netherlands

July 18, 2024

**3 Supplementary Annex C: Reporting standards** **22**

**4 References** **24**

### 1 Supplementary Annex A: Methods

In this section of the supplementary material, we provide detailed information about the different parts of our data-driven analysis. Some subsections deal with the computational framework used, while others relate to data and input parameters. The entire analysis was performed in R (64bit 4.2.1) and the code is available via github (BirgitSollie/HPV-COMPARE).

#### 1.1 Bayesian posterior simulation

A recurring theme in our data-driven analysis involves the use of Bayesian analysis to translate uncertainty about the data into credible intervals for the outcomes. Instead of using fixed parameter estimates, we sampled the parameters from posterior distributions derived from the data.

Our analysis concerns the estimation of expected numbers of events in a cohort of girls and boys invited for HPV vaccination. These numbers were computed according to life-table methodology, based on transitions between health states and to death. The parameters describing these transitions were estimated from population-level data. For practical purposes, all data that we used was divided into age groups and instead of rates we estimated event probabilities, i.e. the probability of the event to occur for an individual in a particular age group. For instance, we computed the probability of a cervical cancer diagnosis in age group $[40, 45)$ conditional on having survived to age 40. The posterior distribution for this parameter was obtained as follows.

Let $X_i$ be a random variable equal to the total number of events in the $i$-th age group with $x_i$ equal to its corresponding observation. Let $n_i$ be equal to the total number of individuals at risk in this age group. We assume $n_i$ to be fixed. Let $\theta_i$ be equal to the event probability, and assume a Beta$(0.5, 0.5)$ prior on $\theta_i$. Under the assumption that events between individuals are independent, $X_i$ follows a binomial distribution with parameters $n_i$ and $\theta_i$. We then see that

$$\begin{aligned}\mathbb{P}(\theta_i|x_i) &\propto \theta^{-0.5}(1-\theta)^{-0.5}\theta^{x_i}(1-\theta)^{n_i-x_i}\\ &= \theta^{x_i-0.5}(1-\theta)^{n_i-x_i-0.5},\end{aligned}$$

which means that the posterior distribution of $\theta_i$ follows a Beta$(x_i + 0.5, n_i - x_i + 0.5)$ distribution.

The number of events of interest in the simulated cohort was subsequently obtained by randomly drawing from the posterior distributions for age-specific transition probabilities, with background mortality considered as a competing event. It is noted that we ignored mortality from other HPV-related cancers as competing events. This approximation greatly facilitates the computation, and is acceptable because the type- and site-specific cancer risks are small. The expected loss in life-years $L_k(a_0)$ due to HPV-related cancers ($k = f$ for females and $k = m$ for males) aged $a_0$ years was calculated as the sum over all HPV-associated tumour sites $j$ (cervical, anal, oropharyngeal, vulvar and vaginal cancer in case $k = f$; oropharyngeal, anal and penile cancer in case $k = m$), as follows:

$$L_k(a_0) = \sum_j p_{k,j} \sum_{a \geq a_0} g_{k,j}(a; a_0)\, l_{k,j}(a)\, da.$$

Here, $p_{k,j}$ denotes the tumour- and possibly sex-specific probability that a cancer is caused by HPV, $g_{k,j}(a; a_0)$ represents the sex-specific population-averaged risk of having cancer $j$ diagnosed in age group $a$ conditional on having survived until age $a_0$, and $l_{k,j}(a)$ is the sex-specific number of life-years lost if HPV-associated cancer $j$ is diagnosed in age-group $a$. We refer to the supplementary information of our previous publications for further elaboration on the computation of loss in (quality-adjusted) life years from HPV-related cancers [1,2].

For estimation of event rates in the absence of HPV vaccination, we performed $B = 1000$ random draws for all parameters and performed cohort simulations by selecting the $b$-th draw for each parameter of interest. This resulted in $B$ different outcomes for each particular number of events, with the mean and variance reflecting the expectation and the uncertainty in the data, respectively.

When projecting the effects of HPV vaccination, we made use of the same random draws, augmented by $B$ random draws for disease-specific HPV genotype attributions (see subsection 1.2), type-specific infection risk reductions by age obtained from our HPV transmission model [3], and the probability that an individual was infected with HPV at a particular age given cancer diagnosis in later life (see subsection 1.3). Differences between vaccination scenarios were calculated for each $b$-th draw of all parameters and summarized afterward. Thus, the differences between bivalent and nonavalent HPV vaccination are estimated conditional on uncertainty in the data.

#### 1.2 HPV genotype attributions

The HPV genotype attributions in CIN2/3 were derived from large Dutch population-based trial data [4,5], and estimated specifically for this study using a previously developed maximum likelihood method that adjusts for multiple infections within subjects [6]. The attributions in cancer were obtained from the literature [7-12]. Table 1 gives an overview of the genotype attributions that we used as input for our analysis.

Table 1: **Genotype attribution in CIN2/3 and cancer**

| | 16 | 18 | 31 | 33 | 39 | 45 | 51 | 52 | 58 |
|---|---|---|---|---|---|---|---|---|---|
| **Screening** | | | | | | | | | |
| CIN2/3, age 30 | 0.5628 | 0.0619 | 0.1144 | 0.0691 | 0.0101 | 0.0274 | 0.0364 | 0.0402 | 0.0360 |
| CIN2/3, age 35+ | 0.4698 | 0.0807 | 0.1317 | 0.0665 | 0.0253 | 0.0233 | 0.0482 | 0.0569 | 0.0427 |
| **Cancer** | | | | | | | | | |
| Cervix | 0.6550 | 0.0729 | 0.0335 | 0.0569 | 0.0131 | 0.0389 | 0.0136 | 0.0194 | 0.0131 |
| Anus | 0.8082 | 0.0365 | 0.0183 | 0.0274 | 0.0046 | 0.0091 | 0.0000 | 0.0068 | 0.0183 |
| Oropharynx | 0.8819 | 0.0176 | 0.0032 | 0.0235 | 0.0016 | 0.0037 | 0.0000 | 0.0016 | 0.0059 |
| Vulva | 0.7283 | 0.0468 | 0.0094 | 0.0656 | 0.0070 | 0.0328 | 0.0000 | 0.0187 | 0.0094 |
| Vagina | 0.5875 | 0.0495 | 0.0528 | 0.0495 | 0.0198 | 0.0363 | 0.0231 | 0.0297 | 0.0363 |
| Penis | 0.6877 | 0.0150 | 0.0090 | 0.0300 | 0.0060 | 0.0270 | 0.0090 | 0.0150 | 0.0120 |

In line with subsection 1.1, we sampled the genotype attributions from posterior distributions of which the mean values are equal to the values in Table 1. In this case, however, the parameter is a vector, $\theta = (\theta_{16}, \theta_{18}, \theta_{31}, \theta_{33}, \theta_{39}, \theta_{45}, \theta_{51}, \theta_{52}, \theta_{58}, \theta_{\text{other}})$, where $\theta_j$ defines the fraction of CIN2/3 lesions (or cancers) in the population caused by HPV genotype $j$ and $\sum_j \theta_j = 1$. Let $X = (X_{16}, X_{18}, X_{31}, X_{33}, X_{39}, X_{45}, X_{51}, X_{52}, X_{58}, X_{\text{other}})$, where $X_j$ is equal to the number of cases in the data caused by HPV type $j$. Let $x = (x_{16}, \ldots, x_{\text{other}})$ be the corresponding vector of observations, with $n = \sum_j x_j$ equal to the total number of cases. We assume $n$ to be fixed and assume a Dirichlet$(0.5, \ldots, 0.5)$ prior on $\theta$. Under the assumption that all individuals are independent, $X$ follows a multinomial distribution with parameters $n$ and $\theta$. Using a similar reasoning as above, it can be shown that the posterior distribution of $\theta$ then follows a Dirichlet$(x + (0.5, \ldots, 0.5))$ distribution.

#### 1.3 From infection risk to cancer incidence reductions

The time between HPV infection and the development of invasive cancer is usually long, possibly decades. Therefore, in the presence of waning efficacy, the probability that an observed cancer case can be prevented by vaccination depends on the age distribution of HPV acquisition. Stated differently, we need a way to translate the age-specific reduction in HPV infection risk, obtained from the HPV transmission model, into reduction in the incidence of HPV-related cancers. In this subsection, we discuss step by step how we achieved this. First, we estimated the time between HPV infection and cancer diagnosis based on the age distributions in the incidence of HPV infection and cancer diagnosis. Second, we calculated the conditional probability distribution of the age of infection given a cancer diagnosis in a particular age group. Third, we projected the reduction in cancer diagnoses by age group based on the age-specific risk reductions for HPV infection.

##### 1.3.1 Setting

Given development of cancer caused by HPV, the age of diagnosis can be modelled as follows. Let $T_1$ be the age of HPV infection and $T_2$ the time of that HPV infection to the cancer diagnosis (given that the infection will develop into cancer). Hence $T_{ca} = T_1 + T_2$ is equal to the age of cancer diagnosis caused by HPV. We assume $T_1$ and $T_2$ to be independent. Note that $T_{ca}$ is only observed when the individual is still alive at time $T_{ca}$.

In the following, we assume that the HPV incidence by age is known from the HPV transmission model (see subsection 1.3.3). The distribution of $T_1$ then follows from this HPV infection incidence by conditioning on acquiring HPV somewhere during life. We discretize time in years resulting in a discrete probability distribution on the sample space $\{1, 2, \ldots, 100\}$ with corresponding probabilities $p_1, p_2, \ldots, p_{100}$ where $\sum_{a=1}^{100} p_a = 1$. We further assume that $T_2$ follows a gamma distribution on $(0, \infty)$ with unknown parameters $k, \theta > 0$ and density function

$$f_{T_2}(t) = \Gamma^{-1}(k)\theta^{-k}t^{k-1}e^{-t/\theta}.$$

In the next subsection we show how the parameters $k$ and $\theta$ can be estimated from the distribution of $T_1$ and cancer incidence data. A key component is that given the distributions of $T_1$ and $T_2$, the distribution of $T_{ca} = T_1 + T_2$ follows from the convolution of the distribution of $T_1$ and the density of $T_2$;

$$f_{T_{ca}}(t) = \sum_a p_a \, f_{T_2}(t - a).$$

##### 1.3.2 Estimation of gamma parameters

Our goal is to estimate the gamma parameters $k$ and $\theta$ in the density of $T_2$ from the distribution of $T_1$ and cancer incidence data. Suppose we have data on the actual number of cancer diagnoses between age $b_1$ and $b_{\ell+1}$, subdivided in $\ell$ different age groups. We define these age groups as $[b_1, b_2)$, $[b_2, b_3)$, $\dots$, $[b_\ell, b_{\ell+1})$. We introduce random variables $X_1, \dots, X_\ell$, where $X_i$ is equal to the number of actual cancer diagnoses in the $i$-th age group. Observed values of $X_1, \dots, X_\ell$ are denoted by $x_1, \dots, x_\ell$. We can estimate $k$ and $\theta$ by likelihood maximization. The likelihood function can be written as

$$\mathcal{L}(k, \theta | X_1 = x_1, \dots, X_\ell = x_\ell) = \mathbb{P}_{k,\theta}(X_1 = x_1, \dots, X_\ell = x_\ell).$$

Given that a cancer is diagnosed, $(X_1, \dots, X_\ell)$ follows a multinomial distribution with probabilities $q_1, \dots, q_\ell$, where $q_i = \mathbb{P}(b_i \le T_{ca} < b_{i+1} | T_{ca} < T_d)$ with $T_{ca}$ equal to the age of cancer diagnosis as defined in the previous subsection and $T_d$ equal to the age of death (by other causes). We assume that $T_{ca}$ and $T_d$ are independent. Then

$$\mathbb{P}_{k,\theta}(X_1 = x_1, \dots, X_\ell = x_\ell) = \frac{\sum_{i=1}^{\ell} x_i}{x_1!, \dots, x_\ell!} q_1^{x_1} \cdots q_\ell^{x_\ell},$$

hence

$$\log \mathcal{L}(k, \theta | X_1 = x_1, \dots, X_\ell = x_\ell) \propto \sum_{i=1}^{\ell} x_i \log q_i. \tag{1}$$

Note that $q_i$ depends on the unknown parameters $k$ and $\theta$, and

$$q_i = \frac{\mathbb{P}(b_i \le T_{ca} < b_{i+1}, T_{ca} < T_d)}{\mathbb{P}(T_{ca} < T_d)}.$$

We have

$$\mathbb{P}(T_{ca} < T_d) = \int_0^\infty f_{T_{ca}}(t)\, s_t \, dt,$$

where $s_t$ is equal to the survival probability $\mathbb{P}(T_d > t)$. Similarly, we have

$$\mathbb{P}(b_i \le T_{ca} < b_{i+1}, T_{ca} < T_d) = \int_{b_i}^{b_{i+1}} f_{T_{ca}}(t)\, s_t \, dt.$$

Hence,

$$q_i = \frac{\int_{b_i}^{b_{i+1}} f_{T_{ca}}(t)\, s_t \, dt}{\int_0^\infty f_{T_{ca}}(t)\, s_t \, dt}. \tag{2}$$

We assume that there are no cancer diagnoses before age $b_1$ and after age $b_{\ell+1}$, so that $\sum q_i = 1$. To evaluate the integrals in (2), we set $s_t = 0$ for $t \ge b_{\ell+1}$. In the previous subsection we have seen how the density of $T_{ca}$ follows from the convolution of $T_1$ and $T_2$. Hence,

$$\begin{aligned}\int_{b_i}^{b_{i+1}} f_{T_{ca}}(t)\, s_t \, dt &= \int_{b_i}^{b_{i+1}} \sum_a p_a \, f_{T_2}(t-a)\, s_t \, dt \\ &= \sum_a p_a \int_{b_i}^{b_{i+1}} f_{T_2}(t-a)\, s_t \, dt.\end{aligned}$$

The survival probabilities are assumed to be available with steps of one year, hence we can write

$$\begin{aligned}\int_{b_i}^{b_{i+1}} f_{T_2}(t-a)\, s_t \, dt &= \sum_{k=1}^{b_{i+1}-b_i} s_{b_i+k-1} \int_{b_i+k-1}^{b_i+k} f_{T_2}(t-a)\, dt \\ &= \sum_{k=1}^{b_{i+1}-b_i} s_{b_i+k-1} \int_{b_i+k-1-a}^{b_i+k-a} f_{T_2}(t)\, dt \\ &= \sum_{k=1}^{b_{i+1}-b_i} s_{b_i+k-1} (F_{T_2}(b_i+k-a) - F_{T_2}(b_i+k-1-a)).\end{aligned}$$

It follows that

$$\int_{b_i}^{b_{i+1}} f_{T_{ca}}(t)\, s_t\, dt = \sum_a p_a \sum_{k=1}^{b_{i+1}-b_i} s_{b_i+k-1}(F_{T_2}(b_i + k - a) - F_{T_2}(b_i + k - 1 - a))$$

and similarly

$$\begin{aligned}\int_0^{\infty} f_{T_{ca}}(t)\, s_t\, dt &= \int_{b_1}^{b_{\ell+1}} f_{T_{ca}}(t)\, s_t\, dt \\ &= \sum_a p_a \sum_{k=1}^{b_{\ell+1}-b_1} s_{b_1+k-1}(F_{T_2}(b_1 + k - a) - F_{T_2}(b_1 + k - 1 - a))\end{aligned}$$

Therefore,

$$q_i = \frac{\sum_a p_a \sum_{k=1}^{b_{i+1}-b_i} s_{b_i+k-1}(F_{T_2}(b_i + k - a) - F_{T_2}(b_i + k - 1 - a))}{\sum_a p_a \sum_{k=1}^{b_{\ell+1}-b_1} s_{b_1+k-1}(F_{T_2}(b_1 + k - a) - F_{T_2}(b_1 + k - 1 - a))}.$$

Using the above expression for $q_i$, the right-hand side of (1) can be maximized numerically (e.g. using the `optim()` function in `R`), which gives the maximum likelihood estimates for $k$ and $\theta$.

##### 1.3.3 Estimated durations to cancer diagnosis

Here, we present pooled estimates of the time from a general high-risk HPV infection to diagnosis of various types of cancer, using cancer incidence data collected from the Netherlands Cancer Registry for the years 2015-2019. This cancer data is given in 15 age groups of width 5 between age 15 and 90 ($b_1 = 15, b_2 = 20, \ldots, b_\ell = 85, b_{\ell+1} = 90$). To approximate the incidence of a general high-risk HPV infection from type-specific HPV infections, we combined the type-specific estimates obtained from the HPV transmission model [3] (in a setting without HPV vaccination) with HPV genotype data from two large population-based screening trials in the Netherlands [4,5]. In these trials, genotyping was assessed for HPV-positive women at baseline for 14 (possibly) high-risk HPV genotypes. We approximated the overall high-risk HPV infection incidence by a weighted sum of the type-specific HPV infection incidences, where the weights are based on the proportion of genotypes found in the data. We used a hierachical classification method. We first ranked the genotypes from most prevalent to least prevalent, resulting in the following ranking; 16, 31, 18, 52, 51, 56, 45, 33, 58, 66, 39, 35, 59, 68. Each genotype was then weighted by the fraction of women positive for that genotype and negative for all genotypes that were more prevalent in the data (ranked higher). We assumed the weights to be the same for men and women.

Figure 1 displays the age distributions of a general HPV infection and diagnosis of various types of cancer used as input for the estimation. Table 2 shows the estimated gamma parameters and the corresponding mean duration from overall high-risk HPV infection incidence to cancer diagnosis. Note that cervical cancer is diagnosed at much younger age then the other cancers, which is also reflected in the mean duration in Table 2 .

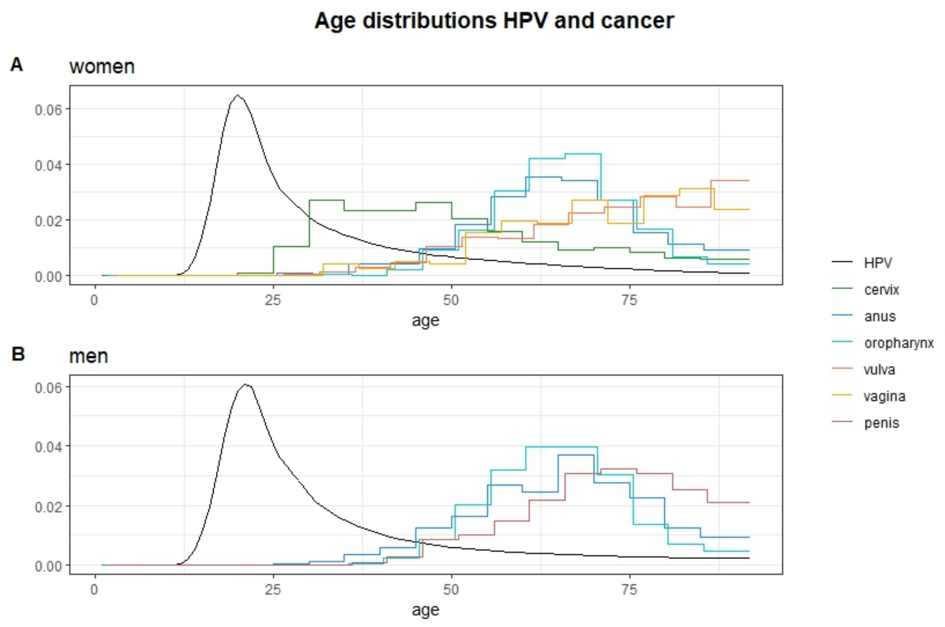

Figure 1: The age distributions for a general HPV infection (black) and cancer, for women in panel A and men in panel B.

Table 2: **Estimated gamma parameters**

| Cancer type | $k$ | $\theta$ | mean duration $(k \cdot \theta)$ |
|---|---|---|---|
| Cervix | 5.39 | 3.62 | 19.54 |
| Anus (w) | 20.91 | 1.91 | 39.97 |
| Anus (m) | 13.09 | 2.92 | 38.26 |
| Oropharynx (w) | 24.78 | 1.58 | 39.18 |
| Oropharynx (m) | 25.73 | 1.47 | 37.82 |
| Vulva | 8.19 | 5.00 | 40.97 |
| Vagina | 10.33 | 3.90 | 40.33 |
| Penis | 22.27 | 1.96 | 43.56 |

##### 1.3.4 Probability of HPV infection age given cancer diagnosis

Our next goal is to compute the conditional probability

$$\mathbb{P}(T_1 = a \,|\, b_i \leq T_{ca} < b_{i+1}, T_{ca} < T_d),$$

that is, the probability that an individual was infected with HPV at age $a$ given that cancer (caused by this infection) was diagnosed in the age interval $[b_i, b_{i+1})$. We will need this probability later to compute risk reductions in cancer. By the definition of a conditional probability we have

$$\mathbb{P}(T_1 = a \,|\, b_i \leq T < b_{i+1}, T_{ca} < T_d) = \frac{\mathbb{P}(T_1 = a, b_i \leq T < b_{i+1}, T_{ca} < T_d)}{\mathbb{P}(b_i \leq T < b_{i+1}, T_{ca} < T_d)}. \tag{3}$$

Remember that $T_{ca} = T_1 + T_2$ and $T_1$, $T_2$ and $T_d$ are independent, hence for the numerator we have

$$\begin{aligned}\mathbb{P}(T_1 = a, b_i \leq T < b_{i+1}, T_{ca} < T_d) &= p_a \int_{b_i - a}^{b_{i+1}-a} f_{T_2}(t)\, s_{t+a}\, dt \\ &= p_a \int_{b_i}^{b_{i+1}} f_{T_2}(t-a)\, s_t\, dt\end{aligned}$$

The denominator follows from the formula of the numerator by summing over all values of $T_1$:

$$\begin{aligned}\mathbb{P}(b_i \leq T < b_{i+1}, T_{ca} < T_d) &= \sum_a \mathbb{P}(T_1 = a, b_i \leq T < b_{i+1}, T_{ca} < T_d) \\ &= \sum_a p_a \int_{b_i}^{b_{i+1}} f_{T_2}(t-a)\, s_t\, dt.\end{aligned}$$

Combining the expressions for the numerator and the denominator in (3), we obtain

$$\mathbb{P}(T_1 = a | b_i \leq T < b_{i+1}, T_{ca} < T_d) = \frac{p_a \int_{b_i}^{b_{i+1}} f_{T_2}(t-a)\, s_t\, dt}{\sum_a p_a \int_{b_i}^{b_{i+1}} f_{T_2}(t-a)\, s_t\, dt}.$$

##### 1.3.5 Number of cancers prevented

In this last part we use the conditional probability $\mathbb{P}(T_1 = a \,|\, b_i \leq T < b_{i+1}, T_{ca} < T_d)$ to compute the risk reduction on cancer. From this point on we need to distinguish between the different HPV types, since the vaccines only protect against a subset of HPV genotypes. We assume that the distribution of $T_1$ and $T_2$ is the same for all types.

Let $RRR_a^j$ be the relative risk reduction on HPV infection with type $j$ at age $a$ corresponding to a chosen vaccination strategy. The risk reductions are estimated by the HPV transmission model and include both direct vaccine effects and indirect herd effects. Then for each age group $[b_i, b_{i+1})$ of cancer diagnosis caused by HPV type $j$, we compute the relative reduction in cancer incidence as

$$RRR_{b_i, b_{i+1}}^{ca,j} = \sum_a \mathbb{P}(T_1 = a \,|\, b_i \leq T < b_{i+1}, T_{ca} < T_d)\, RRR_a^j.$$

To obtain the number of cancers prevented for this age group and HPV type $j$, we multiply $RRR_{b_i, b_{i+1}}^{ca,j}$ with the number of cancers expected for that age group and HPV genotype in the absence of HPV vaccination.

#### 1.4 Anogenital warts episodes

##### 1.4.1 Incidence

Population-level data on anogenital warts (AGW) episodes in the Netherlands was available from general practitioners (GPs), aggregated into very broad age groups [13], see Table 3. It is noted that the incidence rate of AGW is slightly underestimated, because some episodes are diagnosed in sexual health centers, which we did not take into account. However, about 95% of all AGW diagnoses in the Netherlands are made by the GP [13], so the bias is small.

Table 3: **Anogenital warts episodes per 1000 individuals in 2020**

| | $< 25$ | $\geq 25$ |
|---|---|---|
| Women | 2.7 | 2.1 |
| Men | 2.1 | 3.5 |

To obtain fine-grained age trends in AGW episodes from age 25 onward, we used reported trends in the number of sexual partners by 15-year age groups, for men and women [14,15]. Fine-grained trends in AGW episodes below age 25 were obtained from a recent GP registry study, that used a representative sample of about 10% of the Dutch population to estimate the incidence of AGW diagnoses in primary care by age among adolescent girls and young women aged 12-22 years [16]. These rates were re-scaled, with linear interpolation for age 22 to 25, to match the population-averaged incidence rate among women below age 25. For males, we assumed the same age trend as for females aged 12-22 years, and re-scaled results to also match the population-averaged incidence rate among young men below age 25. The resulting estimate of AGW episodes by age is shown in Figure 2.

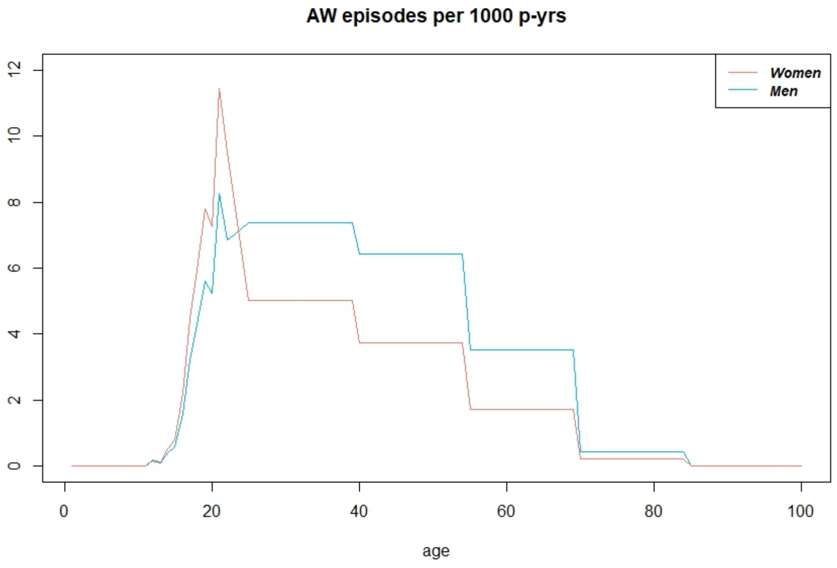

Figure 2: Estimated AGW incidence per 1000 person years (p-yrs) for women (red) and men (blue).

##### 1.4.2 Treatment costs

The costs per treatment episode for anogenital warts were obtained from an epidemiological analysis on the economic burden of genital warts in Dutch primary care [17]. This study used data from the Nivel primary care database over the years 2011-2021. The total costs of treating warts in Dutch primary care increased from EUR 2.3 million in 2011 to EUR 4.9 million in 2021. The costs per case including referrals to secondary care increased from EUR 93.3 in 2015 to EUR 117.4 in 2021. After extrapolation and indexation to 2023, we arrived at treatment costs of EUR 128.7 per anogenital warts episode for use in this analysis.

#### 1.5 Recurrent respiratory papillomatosis

##### 1.5.1 Incidence

We used international publications to obtain the age-specific incidence of recurrent respiratory papillomatosis (RRP). We assumed a prevalence of 1/200 000 person-years for juvenile onset RRP and a prevalence of 1/500 000 person-years for adult onset RRP [18]. For each patient we assumed an exponentially distributed duration with a mean of 10 years. The age distribution of RRP onset was assumed to follow a mixture of three log-normal distributions [19] and is displayed in Figure 3.

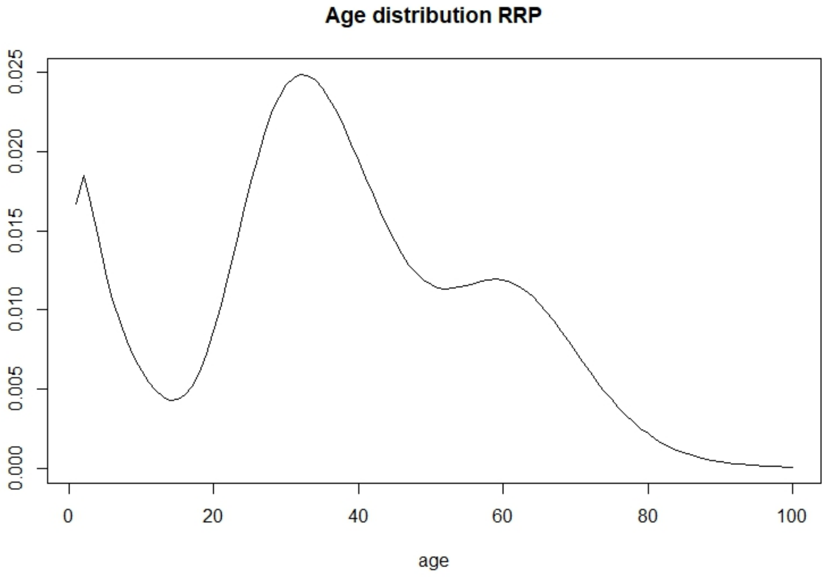

Figure 3: Age distribution of RRP onset

##### 1.5.2 Patient costs

RRP patient costs were calculated per patient per year, using a microcosting approach based on unpublished data from the two academic medical centers in Amsterdam in calendar years 2018 and 2019, with 84 and 97 patients treated in either year, respectively [20]. Taken together, approximately 25% of all RRP patients in the Netherlands are seen in either of these two hospitals.

The direct costs included outpatient visit costs, day care and hospitalization costs, diagnostic costs (including laryngology, morphology of biopsy samples, ultrasound, blood tests), and therapeutic laryngology costs. Unit direct costs were based on tariffs used in the two hospitals in 2018 and 2019. The indirect costs included production loss costs and travel costs to the academic medical center. Production loss costs per hour and travel costs per kilometer were taken from [21]. Distance to the hospital was calculated from zip codes of the patient's home address and the hospital. Travel costs comprised 10% and 13% of the indirect costs in years 2018 and 2019, respectively. The production loss costs were adjusted for labor participation percentages, stratified by age and sex, as reported by Statistics Netherlands. Production loss costs also included costs of assisting children with RRP to the hospital.

Average direct and indirect costs per patient per year, weighted by the number of patients in the two hospitals, are presented in Table 4. Summing the average direct and indirect costs yielded a total of EUR 2579 per RRP patient per year.

Table 4: **Costs per RRP patient per year**

| **Direct costs** | Cost (€) | **Indirect costs** | Cost (€) |
|---|---|---|---|
| Outpatient consult visit | 249.30 | Production loss | 510.30 |
| Diagnostics | 246.40 | Travel costs | 51.70 |
| Treatment (laryngoscopy) | 1044.10 | | |
| Daycare / hospitalization | 313.70 | | |
| Other (e.g. speech therapy) | 118.10 | | |

All costs were indexed to the year 2023.

#### 1.6 Cancer screening and treatment costs

Costs of cancer screening and treatment are presented in Table 1 of the main article. Costs of cancer treatment are based on a previous publication [22], and updated to 2023 using the consumer price index. Costs of screening, including colposcopy referral following a positive HPV-test and non-normal cytology, were based on the subsidy regulation to the cervical cancer screening program determined by the Dutch government. The costs of follow-up and treatment of CIN2/3 is elaborated below.

##### 1.6.1 CIN2/3 patient costs

The costs per CIN2/3 diagnosis were calculated from the nationwide network and registry of histo- and cytopathology [23] and a population-based screening trial database [24]. The CIN2/3 costs included the number of indicative screening tests after colposcopy referral but before diagnosis, the number of follow-up screening tests after treatment (either cytology or HPV-testing), and the number of colposcopy-guided biopsies. We assumed that all CIN3 cases and 65% of CIN2 cases were eventually treated and that treatment of residual CIN2/3 was needed in 15% of the cases [23,25,26]. Unit costs for the indicative and follow-up screening tests were taken from [27], unit LLETZ treatment costs were taken from [28], and costs per outpatient clinic visit were taken from [21]. The number of procedures per patient and costs per CIN2 and CIN3 diagnosis are presented in Table 5.

Table 5: **Direct costs per CIN2/3 diagnosis**

| | Cost category | No. | Cost per procedure (€) | Cost per diagnosis (€) | Source |
|---|---|---|---|---|---|
| **CIN2** | | | | | |
| | Colposcopies | 4.56 | 191.70 (first) | 757 | [21,25] |
| | | | 158.80 (repeat) | | |
| | Biopsies | 2.01 | 70.10 | 141 | [21,25] |
| | LLETZ | 0.75 | 701.00 | 526 | [19-21,23,25] |
| | Co-tests after treatment | 2.55 | 60.60 | 155 | [21,25] |
| **Total** | | | | **1578** | |
| **CIN3** | | | | | |
| | Colposcopies | 4.89 | 191.70 (first) | 809 | [21,25] |
| | | | 158.80 (repeat) | | |
| | Biopsies | 2.30 | 70.10 | 161 | [21,25] |
| | LLETZ | 1.15 | 701.00 | 806 | [19-21,23,25] |
| | Co-tests after treatment | 2.59 | 60.60 | 157 | [21,25] |
| **Total** | | | | **1934** | |

All costs were indexed to the year 2023.

#### 1.7 Utilities for non-lethal conditions

In sensitivity analyses, we also expressed the incremental cost-effectiveness ratio (ICER) of 9vHPV versus 2vHPV vaccination in terms of the cost per quality-adjusted life-years (QALYs) gained. This acknowledges that 9vHPV confers additional health benefits by not only reducing the incidence of HPV-related cancers relative to 2vHPV vaccination, but also by reducing the occurrence of non-lethal conditions. To this end, we attributed a loss in quality of life to precancerous lesions, anogenital warts and RRP. For precancerous lesions, we assumed a QALY loss of 0.035 per CIN2/3 detected [29]. For anogenital warts, we attributed a QALY loss of 0.018 for each treatment episode [30]. For RRP, we assumed a QALY loss of 0.105 per patient per year [31].

#### 2 Supplementary Annex B: Results

In this section of the supplementary material we provide more numerical and graphical results.

##### 2.1 Sensitivity analysis

Table 6: **Quantiles of ICERs of 9vHPV versus 2vHPV vaccination for different scenarios corresponding to the one-way and two-way sensitivity analyses**

| Scenario | $Q_{0.025}$ | $Q_{0.25}$ | $Q_{0.5}$ | $Q_{0.75}$ | $Q_{0.975}$ |
|---|---|---|---|---|---|
| **One-way** | | | | | |
| Base-case | 4166 | 5588 | 6192 | 6758 | 7916 |
| No LR types | 14719 | 16389 | 17173 | 18060 | 19866 |
| LR elimination | 1941 | 3494 | 4120 | 4701 | 5824 |
| No cross-protection | -2259 | -1776 | -1566 | -1348 | -973 |
| High cross-protection | 9300 | 11776 | 12882 | 14081 | 16589 |
| Very high cross-protection | 16040 | 20181 | 22558 | 25037 | 32709 |
| QALYs instead of LYs | 1412 | 1821 | 2009 | 2201 | 2514 |
| HPV vaccine uptake 50% | 2409 | 3600 | 4114 | 4634 | 5697 |
| HPV vaccine uptake 70% | 10438 | 11954 | 12888 | 13758 | 15631 |
| Price difference EUR 35 | -3484 | -2346 | -1830 | -1321 | -534 |
| Price difference EUR 70 | 13949 | 16004 | 16839 | 17810 | 19922 |
| Waning | 48995 | 56057 | 59499 | 63788 | 72315 |
| International discounting | 7614 | 10209 | 11297 | 12362 | 14565 |
| **Two-way** | | | | | |
| Very high cross-protection + LR elimination | 12267 | 16060 | 18163 | 20490 | 27120 |
| High cross-protection + no LR types | 25846 | 29259 | 31297 | 33089 | 37435 |
| QALYs instead of LYs + very high cross-protection | 3485 | 4030 | 4322 | 4598 | 5125 |
| HPV vaccine uptake 50% + very high cross-protection | 11046 | 14075 | 15801 | 17853 | 22452 |
| HPV vaccine uptake 70% + no LR types | 21785 | 24010 | 25438 | 26756 | 29551 |
| HPV vaccine uptake 70% + high cross-protection | 24322 | 28514 | 31069 | 33913 | 41720 |
| Price difference EUR 35 + very high cross-protection | 1919 | 4507 | 5750 | 7163 | 10044 |
| Price difference EUR 70 + no LR types | 24041 | 26637 | 27944 | 29225 | 32369 |
| Price difference EUR 70 + high cross-protection | 24828 | 28658 | 30740 | 32811 | 37457 |
| Price difference EUR 70 + HPV vaccine uptake 70% | 23127 | 25550 | 27116 | 28674 | 31826 |
| Waning + LR elimination | 43446 | 50086 | 53364 | 57417 | 65058 |
| Waning + no cross-protection | 18927 | 21185 | 22563 | 23951 | 26772 |
| Waning + QALYs instead of LYs | 7810 | 8573 | 8974 | 9336 | 9941 |
| Waning + HPV vaccine uptake 50% | 40581 | 46610 | 49614 | 53105 | 60611 |
| Waning + Price difference EUR 35 | 28667 | 33513 | 36103 | 38725 | 44547 |

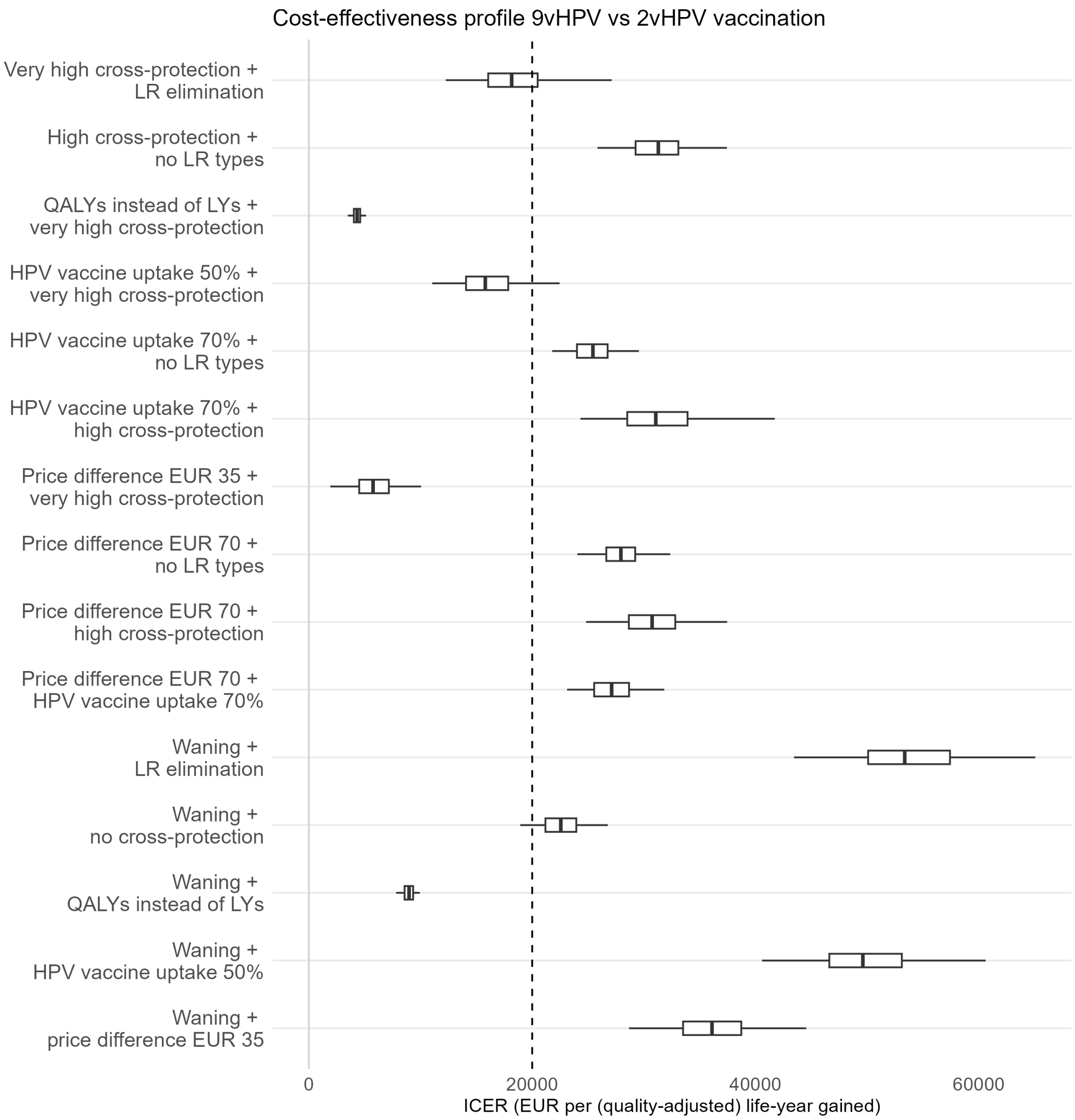

Figure 4: ICERs of 9vHPV versus 2vHPV vaccination for different scenarios in the two-way sensitivity analysis. The light-grey vertical line corresponds to an ICER equal to zero. The cost-effectiveness threshold of €20 000 per life-year gained is displayed by the dashed vertical line. Boxplots display median and interquartile range of predictions, with whiskers denoting the 95% credible intervals

The plots below show the cost-effectiveness planes for the comparison of 9vHPV versus 2vHPV vaccination for various assumptions also considered in the sensitivity analysis. Each plot shows twelve scenarios regarding cross-protection (cp) and evaluated impact on the low-risk HPV types.

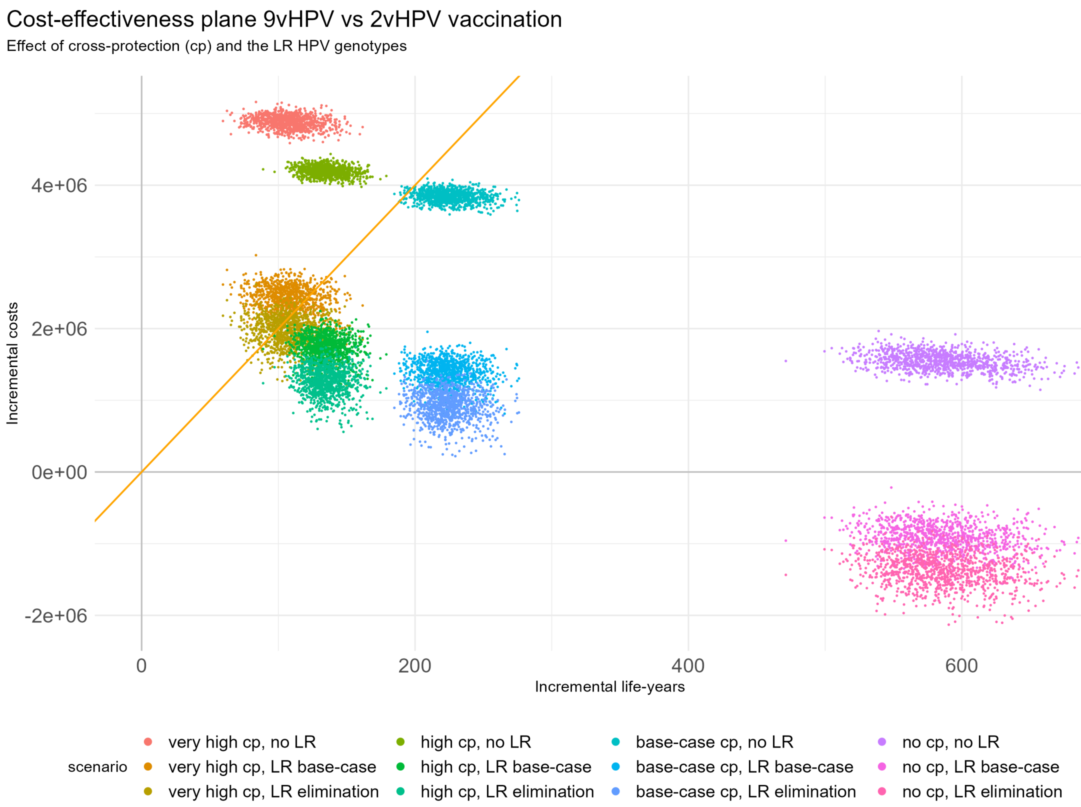

Figure 5: Cost-effectiveness plane of 9vHPV vs 2vHPV vaccination. The cost-effectiveness threshold of €20 000 per life-year gained (LYG) is shown by the orange line.

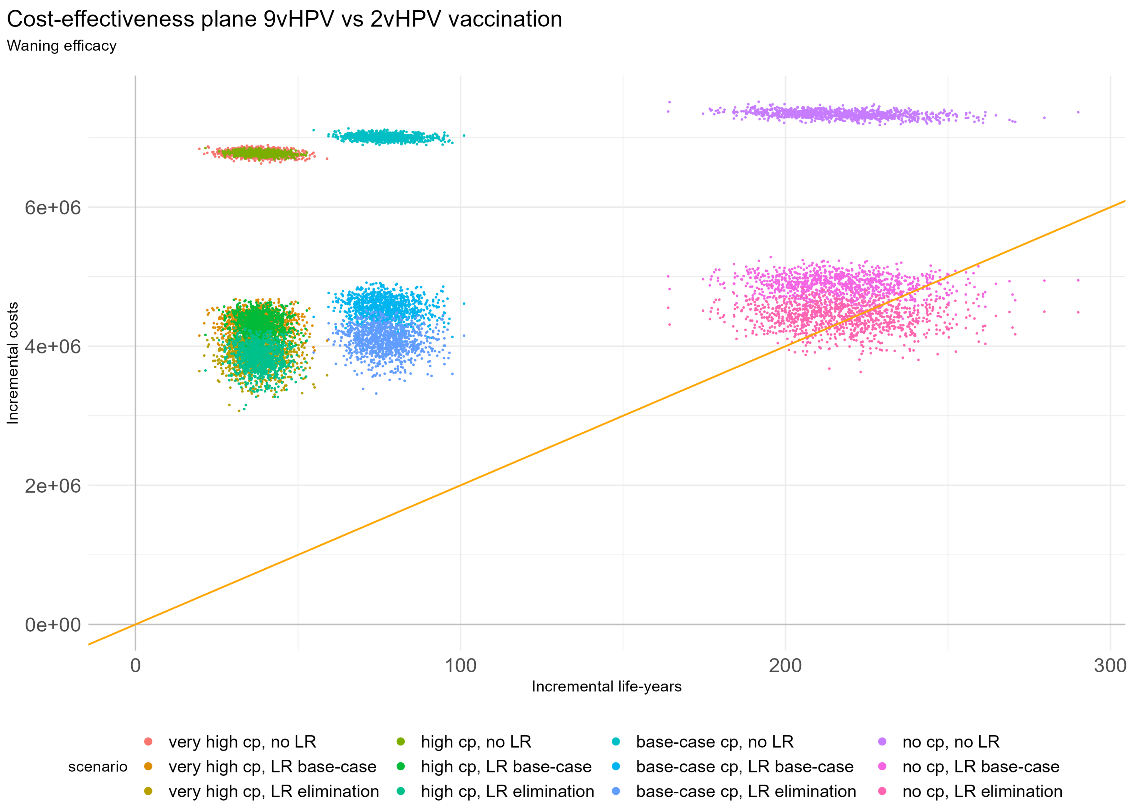

Figure 6: Cost-effectiveness plane of 9vHPV vs 2vHPV vaccination under waning vaccine efficacy. The cost-effectiveness threshold of €20 000 per LYG is shown by the orange line.

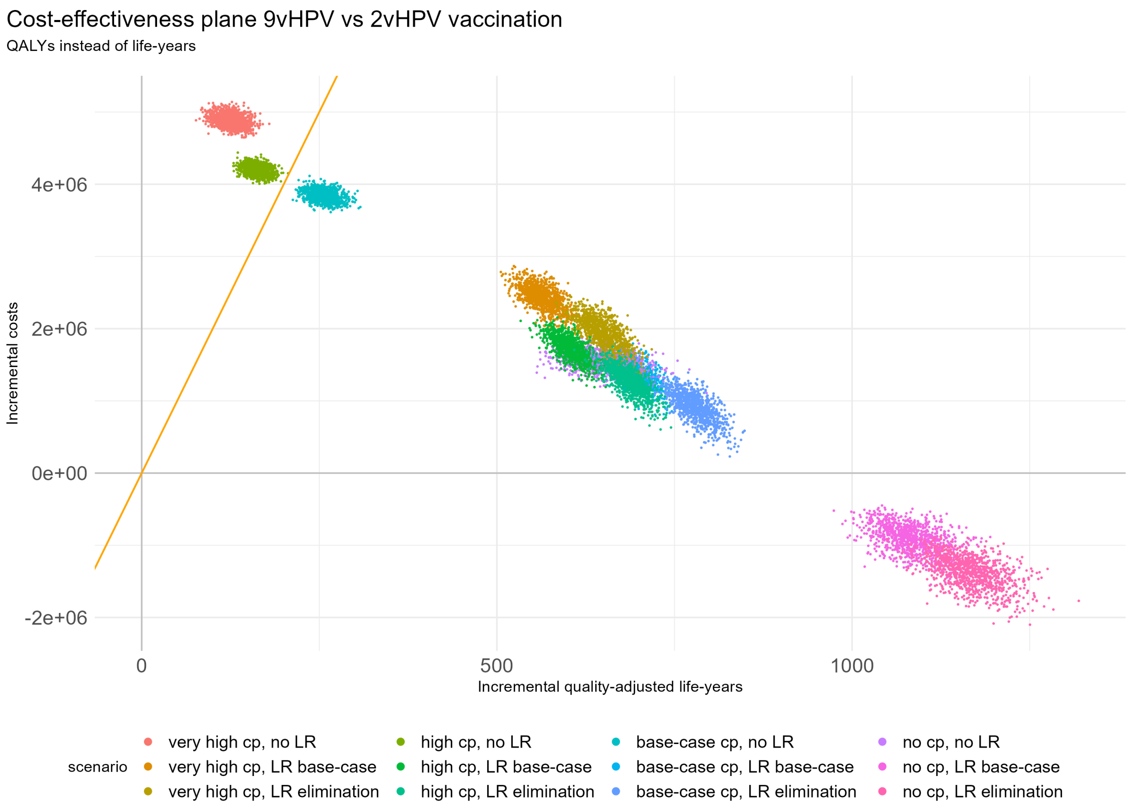

Figure 7: Cost-effectiveness plane of 9vHPV vs 2vHPV vaccination using quality-adjusted life-years gained. The cost-effectiveness threshold of €20 000 per LYG is shown by the orange line.

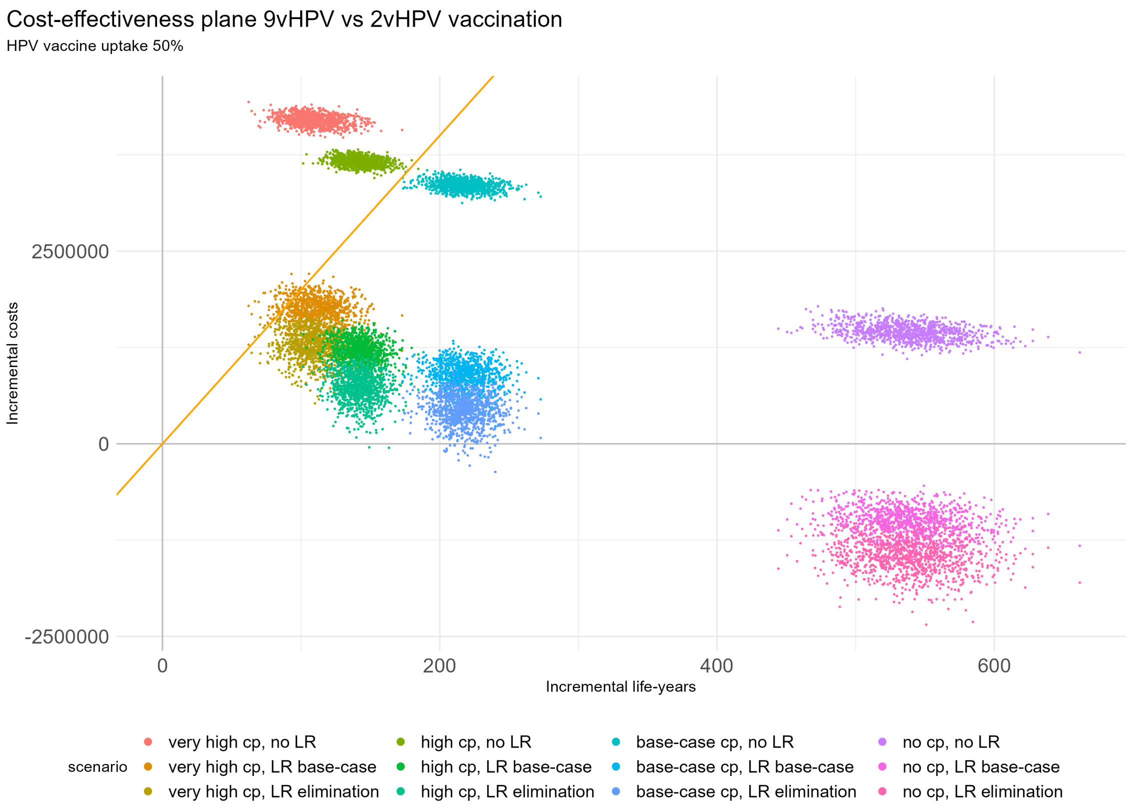

Figure 8: Cost-effectiveness plane of 9vHPV vs 2vHPV vaccination when the vaccine uptake is equal to 50% for both boys and girls. The cost-effectiveness threshold of €20 000 per LYG is shown by the orange line.

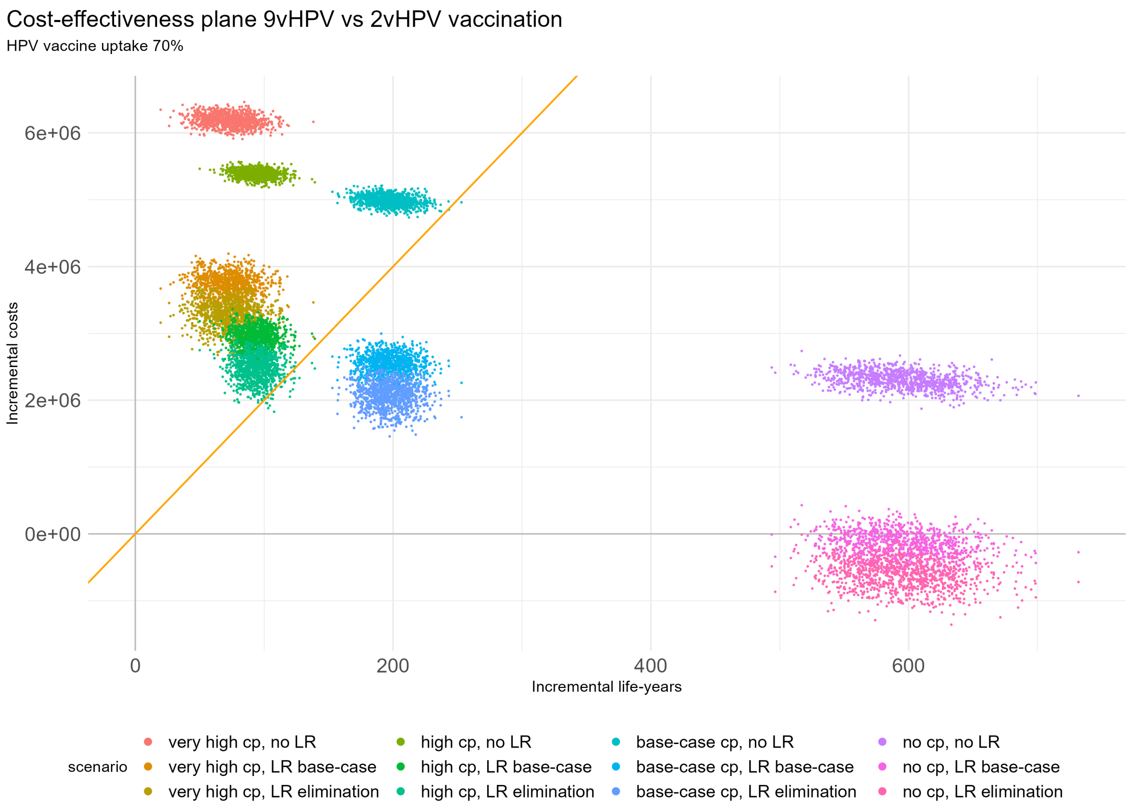

Figure 9: Cost-effectiveness plane of 9vHPV vs 2vHPV vaccination when the vaccine uptake is equal to 70% for both boys and girls. The cost-effectiveness threshold of €20 000 per LYG is shown by the orange line.

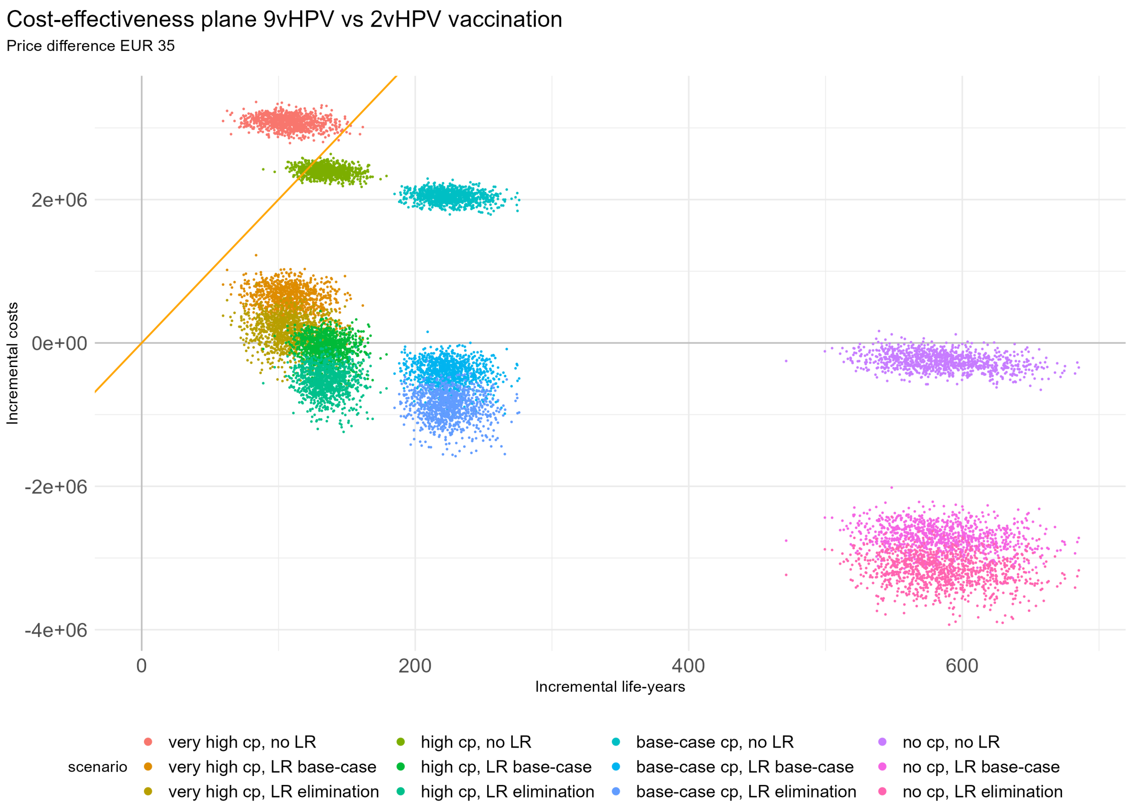

Figure 10: Cost-effectiveness plane of 9vHPV vs 2vHPV vaccination when the price difference between the 9vHPV and 2vHPV vaccines, for a two-dose vaccination schedule, is EUR 35. The cost-effectiveness threshold of €20 000 per LYG is shown by the orange line.

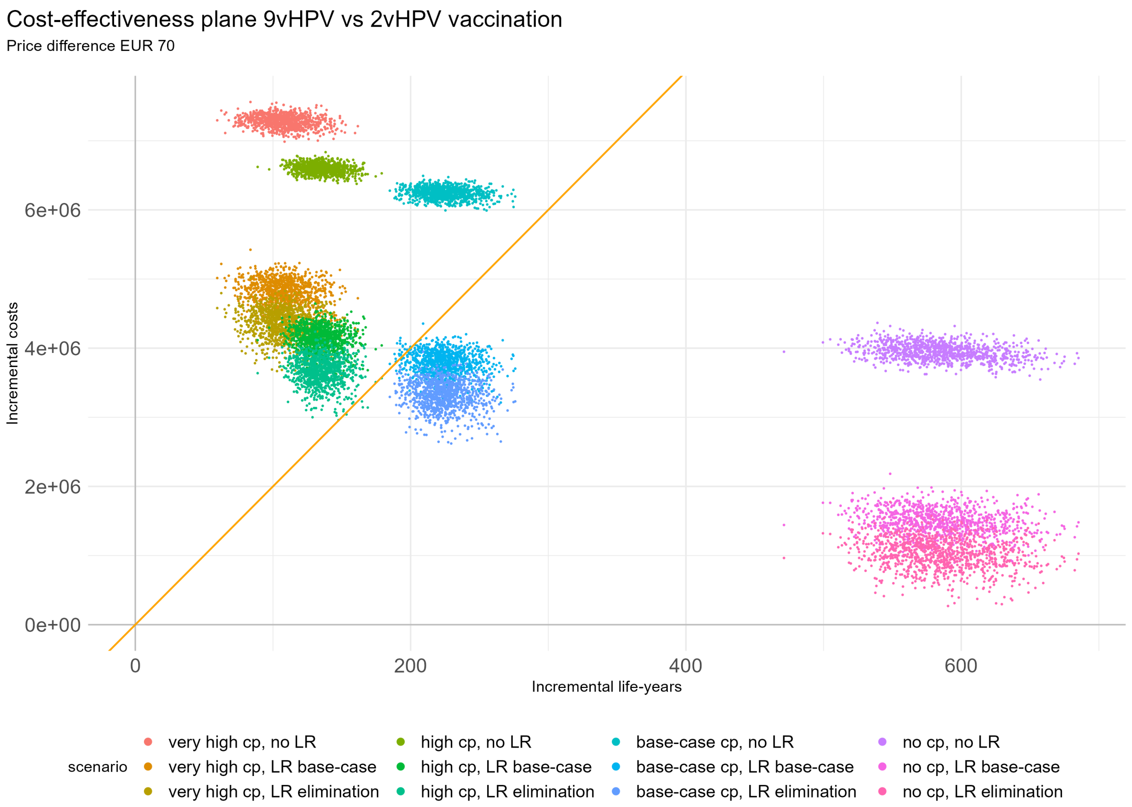

Figure 11: Cost-effectiveness plane of 9vHPV vs 2vHPV vaccination when the price difference between the 9vHPV and 2vHPV vaccines, for a two-dose vaccination schedule, is EUR 70. The cost-effectiveness threshold of €20 000 per LYG is shown by the orange line.

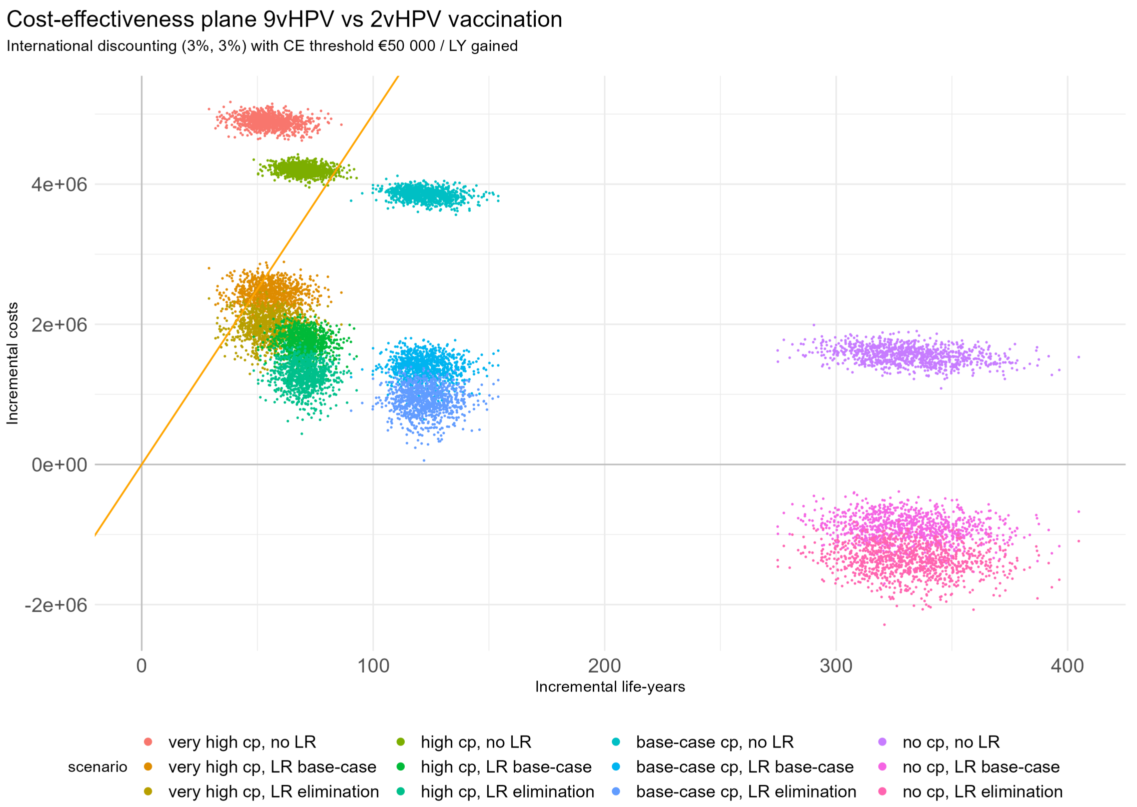

Figure 12: Cost-effectiveness plane of 9vHPV vs 2vHPV vaccination under international discounting of 3% per year for both effects and costs. The cost-effectiveness threshold of €50 000 per LYG is shown by the orange line.

### 3 Supplementary Annex C: Reporting standards

HPV-FRAME is a CONSORT-style itemised checklist encapsulating agreed reporting standards that has been structured according to 7 domains reflecting distinct policy questions in HPV and cancer prevention. Its goal is to support understanding regarding a model's strengths and weaknesses and contribute to equitable, evidence-based decision-making. This investigation adheres to the quality framework for modelled evaluations of HPV-related cancer control through prophylactic vaccination in preadolescence (domain no. 1), for which the core set plus 10 additional reporting standards apply [32]. These are summarized in Table 7.

Table 7: **HPV-FRAME reporting standards for vaccination in (pre)adolescence**

| a) Inputs | Report by age | Report by sex | Comments |
|---|---|---|---|
| Target population for intervention | Y | Y | Vaccination of girls + boys in the Netherlands at age 10 |
| Sexual behaviour | Y (implicitly) | Y (implicitly) | Rates and mixing patterns in original publication [3] |
| Time horizon | Y | Y | Lifetime cohort evaluation at post-vaccination equilibrium |
| Quality of life assumptions | Y | Y | Part of sensitivity analysis |
| Calibration | N/A | N/A | Data-driven analysis, does not rely on calibration |
| Validation | N/A | N/A | No targets available |
| Costs | Y | Y | See Supplementary Annex A |
| Vaccine uptake | Y | Y | Varied in sensitivity analysis, assumed equal by sex |
| Vaccine efficacy | Y | Y | Efficacy informed by CIN2/3, assumed equal for other diseases |
| Vaccine cross-protection | Y | Y | Cross-protection for 2vHPV vaccine, assumed equal by sex |
| Duration vaccine protection & waning | Y | Y | Varied in sensitivity analysis, assumed equal by sex |
| Vaccine delivery and costs | Y | Y | Total costs per two-dose schedule, assumed equal by sex |
| Pre-vaccination disease burden (incl. attributable fractions for HPV) | Y | Y | HPV attributions independent of age, except for CIN2/3 |
| Duration of natural immunity | Y | Y | Exponential waning rates, assumed equal by sex |
| **b) Outputs** | **Reported** | **Report by sex** | **Comments** |
| Absolute reductions in HPV or warts | Y | N | Reductions in anogenital warts and RRP for 9vHPV vaccine |
| Absolute reductions in CIN2/3 | Y | N/A | Reduction in CIN2/3 given for direct and total effects |
| Absolute reductions in invasive cancer | Y | Y | Reductions in cancers given for direct and total effects |

20. Gielgens T, Kliffen T, Berkhof J, Dikkers FG. Annual and direct costs of recurrent respiratory papillomavirus in the Netherlands. Amsterdam UMC; unpublished report.

21. Anonymous. Richtlijn voor het uitvoeren van economische evaluaties in de gezondheidszorg [in Dutch]. Zorginstituut Nederland; 2016.

22. de Kok IM HJ, van Rosmalen J, van Ballegooijen M. Would the effect of HPV vaccination on non-cervical HPV-positive cancers make the difference for its cost-effectiveness? Eur J Cancer. 2011; 47(3):428-35.

23. Anonymous. Verbetersignalement Baarmoederhalsafwijking CIN [in Dutch]. Zinnige Zorg; 2019.

24. Polman NJ, Ebisch RMF, Heideman DAM, Melchers WJG, Bekkers RLM, Molijn AC, et al. Performance of human papillomavirus testing on self-collected versus clinician-collected samples for the detection of cervical intraepithelial neoplasia of grade 2 or worse: a randomised, paired screen-positive, non-inferiority trial. Lancet Oncol. 2019; 20(2):229-238.

25. Kremer WW, Dick S, Heideman DAM, Steenbergen RDM, Bleeker MCG, Verhoeve HR, et al. Clinical regression of high-grade cervical intraepithelial neoplasia is associated With absence of

FAM19A4/miR124-2 DNA methylation (CONCERVE Study). J Clin Oncol. 2022; 40(26):3037-46.

26. Kocken M, Helmerhorst TJ, Berkhof J, Louwers JA, Nobbenhuis MA, Bais AG, et al. Risk of recurrent high-grade cervical intraepithelial neoplasia after successful treatment: a long-term multi-cohort study. Lancet Oncol. 2011; 12(5):441-50.

27. Jansen E, Naber SK, Aitken CA, de Koning HJ, van Ballegooijen M, de Kok I. Cost-effectiveness of HPV-based cervical screening based on first year results in the Netherlands: a modelling study. BJOG. 2021; 128(3):573-82.

28. van Ballegooijen M, Rebolj M, Essink-Bot ML, Meerding WJ, Berkers LM, Habbema JDF. De effecten en kosten van het bevolkingsonderzoek naar baarmoederhalskanker in Nederland na de herstructurering [in Dutch]. Erasmus MC, Rotterdam; 2006.

29. Berkhof J, Bogaards JA, Demirel E, Diaz M, Sharma M, Kim JJ. Cost-effectiveness of cervical cancer prevention in Central and Eastern Europe and Central Asia. Vaccine. 2013; 31(Suppl 7):H71-9.

30. Jit M, Chapman R, Hughes O, Choi YH. Comparing bivalent and quadrivalent human papillomavirus vaccines: economic evaluation based on transmission model. BMJ. 2011; 343:d5775.

31. Chesson HW, Forhan SE, Gottlieb SL, Markowitz LE. The potential health and economic benefits of preventing recurrent respiratory papillomatosis through quadrivalent human papillomavirus vaccination. Vaccine. 2008; 26(35):4513-8.

32. Canfell K, Kim JJ, Kulasingam S, Berkhof J, Barnabas R, Bogaards JA, et al. HPV-FRAME: A consensus statement and quality framework for modelled evaluations of HPV-related cancer control. Papillomavirus Res. 2019; 8:100184.
